# Forecasting regional-level COVID-19 hospitalisation in England as an ordinal variable using the machine learning method

**DOI:** 10.1101/2023.10.17.23297138

**Authors:** Haowei Wang, Kin On Kwok, Ruiyun Li, Steven Riley

**Author notes:** Corresponding authors: Steven Riley, School of Public Health, Imperial College London, Norfolk Place, London, W2 1PG.

## Abstract

**Background:** COVID-19 causes substantial pressure on healthcare, with many healthcare systems now needing to prepare for and mitigate the consequences of surges in demand caused by multiple overlapping waves of infections. Therefore, public health agencies and health system managers also now benefit from short-term forecasts for respiratory infections that allow them to manage services better. However, the availability of easily implemented effective tools for generating precise forecasts at the individual regional level still needs to be improved.

**Methods:** We extended prior work on influenza to forecast regional COVID-19 hospitalisations in England for the period from 19th March 2020 to 31st December 2022, treating the number of hospital admissions in each region as an ordinal variable. We further developed the XGBoost model used previously to forecast influenza to enable it to exploit the ordering information in ordinal hospital admission levels. We incorporated different types of data as predictors: epidemiological data including weekly region COVID-19 cases and hospital admissions, weather conditions and mobility data for multiple categories of locations (e.g., parks, workplaces, etc). The impact of different discretisation methods and the number of ordinal levels was also considered.

**Results:** We find that the inclusion of weather data consistently increases the accuracy of our forecasts compared with models that rely only on the intrinsic epidemiological data, but only by a small amount. Mobility data brings about a more substantial increase in our forecasts. When both weather and mobility data are used in addition to the epidemiological data, the results are very similar to the model with only epidemiological data and mobility data.

**Conclusion:** Accurate ordinal forecasts of COVID-19 hospitalisations can be obtained using XGBoost and mobility data. While uniform ordinal levels show higher apparent accuracy, we recommend N-tile ordinal levels which contain far richer information.

**Author Summary:** At the regional level, we address the pressing need for precise short-term forecasts of respiratory infections, particularly COVID-19. We focus on the specific context of England and cover the period from January 1 to December 31, 2022. We introduced an enhanced XGBoost model that leverages the ordinal nature of hospital admission data, utilising a combination of predictors, including epidemiological data, weather conditions, and mobility data across various location categories. Our findings indicate that the inclusion of weather data marginally improves forecasting accuracy, while mobility data yields more significant enhancements. This research contributes valuable insights for public health agencies and healthcare system managers in their ongoing efforts to manage and respond to the complexities of the COVID-19 pandemic.

## Introduction

The COVID-19 pandemic placed a considerable strain on hospitals at varying times and in different locations between January 2020 and late 2021. By 31st December 2022, the World Health Organization (WHO) has reported more than 600 million confirmed cases, including 6.6 million deaths [1]. Given the significant variation in disease incidence across both space and time, even within the same country, local public health authorities often faced challenges in obtaining adequate insights to effectively prioritise health services. This deficiency led to even greater disruption in healthcare delivery than would have been otherwise anticipated. Having advanced and accurate knowledge of higher disease incidence allowed for the postponement of elective care, whereas insight into lower disease incidence facilitated a more expedited refocusing on the backlog of postponed elective procedures. Heightened levels of uncertainty were especially pronounced during the emergence of new COVID-19 variants, which were associated with increased transmission rates and, at times, greater severity.

Specifically, in England and Wales, there were repeated rapid surges in SARS-CoV-2 infections, resulting in high demands for hospitalisations and medical resources for COVID-19 patients. While vaccination efforts have significantly reduced the need for additional interventions to prevent overwhelming hospitals, even as of December 2022, the persistently high rates of infections and mortality have placed substantial pressure on the resources of the National Health Service (NHS). Consequently, the need for frequently updated short-term forecasting at the local level is evident, as it has the potential to markedly enhance the efficiency with which limited healthcare capacity is employed [2].

Non-epidemiological time-varying factors may have collectively influenced the transmission dynamics of SARS-CoV-2 [3,4]. Numerous studies have detailed the association between COVID-19 cases and climate or meteorological conditions [5,6]. Multiple investigations conducted in France have confirmed that the integration of weather factors into models can enhance the model’s capability to reproduce observed patterns, encompassing the progression of hospital admissions [7,8].

Changes in human social behaviours swiftly impact the trajectory of an epidemic and the influence of severe disease [9,10]. Comprehensive mobility data can be harnessed to illustrate shifts in customary commuting behaviours, patterns of social interaction, and inter-regional transits. These changes in metrics have shown correlations with infection rates and mortality [11]. In this work, we endeavour to expand upon these findings by incorporating weather conditions and mobility patterns into a model designed to predict COVID-19 admissions at the regional level.

Our previous work involved the development of a machine learning short-term forecasting technique which demonstrated success through retrospective evaluations in predicting the incidence of influenza-like illness (ILI) as an ordinal variable [12]. The method exhibited satisfactory performance in these retrospective assessments. In this study, we expand upon our previous method by integrating local weather conditions and mobility patterns to forecast short-term, weekly COVID-19 admissions for seven NHS regions of England. As such, we conduct a two-step evaluation of this novel method: 1) We examine whether the inclusion of weather and mobility data enhances the accuracy of 1-to-4-week forecast individually and collectively drawing from previous admission patterns; 2) We explore whether the machine learning models can consistently achieve high accuracy across various levels of application.

## Methods

### Data

Our dataset amalgamates information encompassing epidemiological aspects of COVID-19 (hospital admissions, cases and deaths), human mobility patterns and weather data. All data were sourced from publicly available outlets. Epidemiological data were publicly available on the GOV.UK Coronavirus (COVID-19) in the dashboard [13]. This source provided daily hospitalisation data for NHS regions in England, capturing instances of positive COVID-19 tests within 14 days prior to hospitalisation, along with post-admission positive cases. However, for NHS regions, daily cases and death statistics were unavailable. Instead, we extracted case and death data for Lower Tier Local Authorities (LTLAs) and subsequently matched the data of 315 LTLAs in England with their respective NHS regions. This facilitated the aggregation of NHS region-specific-case and death data. Daily death data recorded individuals who died within 28 days of being confirmed COVID-19 cases, with the date reflecting the date of death instead of the date of reporting. Hospital admission data became available on 19th March 2020 which postdates the availability of case and death data. To ensure temporal consistency, the analysis encompassed epidemiological data spanning from 19th March 2020 to 31st December 2022.

Daily hospitalisation, cases and deaths were aggregated by ISO week. Subsequently, the weekly numbers were adjusted relative to the regional population, scaled per 100000 people, thereby generating per capita hospitalisation, cases and death statistics. The regional population was deduced from population estimates at the LTLA levels by the Office for National Statistics (ONS) [14]. Per capita cases and deaths remained as continuous variables while per capita hospitalisation was converted into an ordinal variable using distinct methodologies described below.

Per capita hospitalisation was converted into an ordinal variable with numerical levels through two techniques: 1) N-tile, and 2) n-uniform interval, where n = 3, 5, and 10. The N-tile method constitutes an unsupervised discretisation method that segments the value range into a specified number of bins (i.e., 3, 5 and 10 in this context), aiming to maintain nearly uniform instances within each bin This effectively translates numeric target values into ordinal quantities by ensuring equivalent-frequency binning. On the other hand, the n-uniform interval strategy divides the range of values into equidistant bins, each containing varying observation quantities. For both methodologies, the higher the value of the level, the higher the number of hospital admissions, with level 1 representing minimal hospitalisation and level n indicating the highest level of hospitalisation.

We collected human mobility data from Google, which provides aggregated anonymised information sourced from its online platform [15]. This data provides insights into the percentage changes in mobility across 7 different location categories, serving as a measure of movement trends in response to the pandemic-related lockdowns. The reference point for these trends is the baseline day, defined as the median value from 3rd January to 6th February 2020. Each location category has its own specific baseline day. Google stopped reporting new data on 15th October 2022.

While Google provides mobility data at the sub-regional levels for England, denoted by *sub_region_1* and *sub_region_2* columns in the raw data, it is not readily compatible with NHS regions. To address this, we used a combination of *sub_region_1* and *sub_region_2* columns to map LTLA using a lookup table available on the GitHub repository “*datasciencecampus / google-mobility-reports-data” [16]*. By calculating the mean daily mobility values at the LTLA level and associating them with different NHS regions, we were able to derive the daily mobility data for NHS regions. Finally, we obtained weekly mobility data at the NHS-regional level by averaging the daily mobility by ISO weeks.

Our analysis also included weekly weather data, specifically air temperature at 2 meters (m) and total precipitation, obtained from the fifth-generation reanalysis (ERA5) by the European Centre for Medium-Range Weather Forecasts (ECMWF) [17]. To acquire data at the NHS-regional level, we identified the nearest grid point to the centre of each of the 315 LTLAs and calculated weekly temperature and total precipitation averages for the respective NHS regions. Data types and predictors were summarised in Table 1.

**Table 1.**
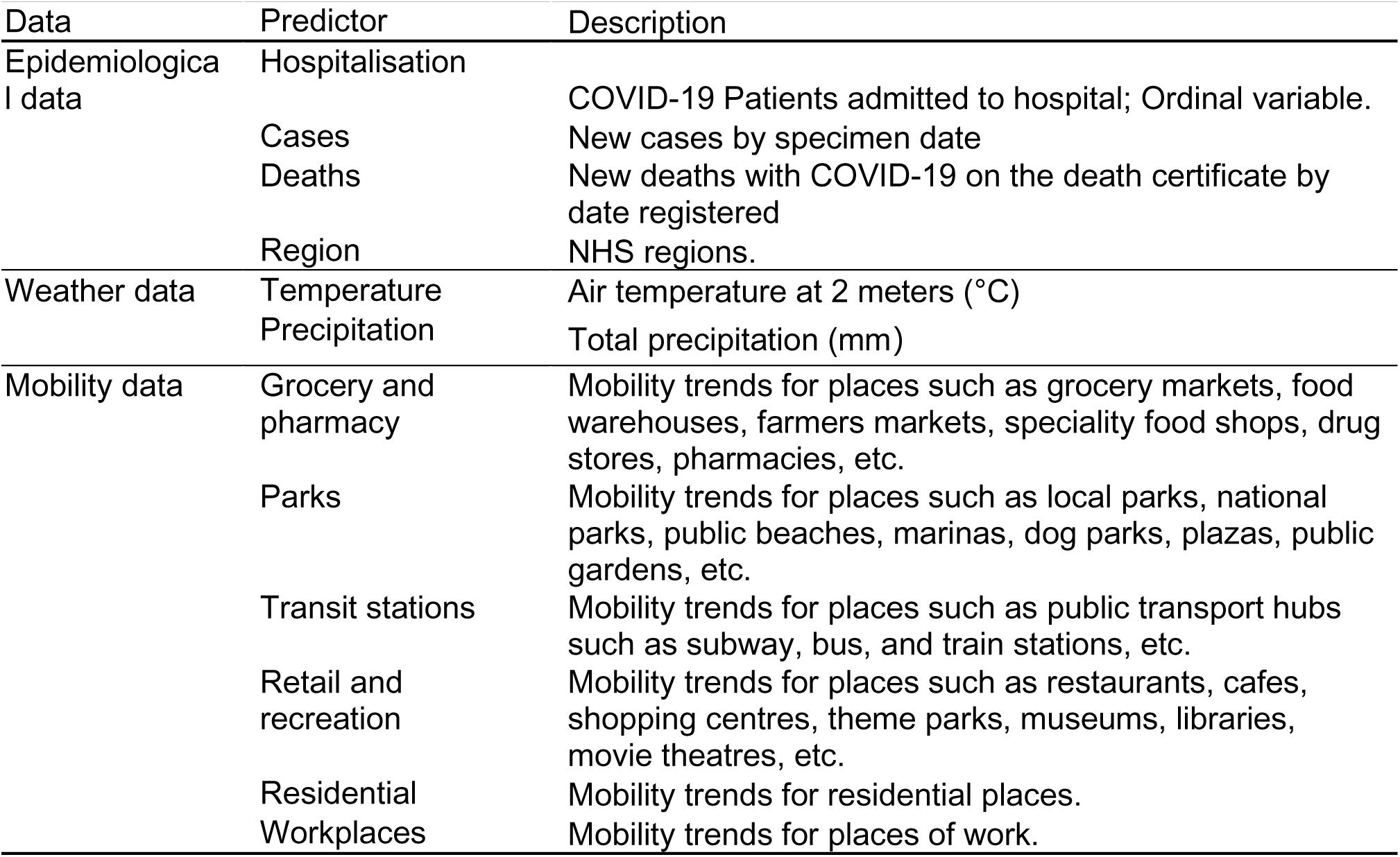
Detailed descriptions of predictors.

### Primary forecast Model

XGBoost is a standard classification algorithm for nominal classes, the ordinal information in the class attributes may be discarded partly when applied to ordinal prediction problems, while information can potentially enhance the predictability of the classifier. Therefore, we used a simple method developed by [18] to enable the underlying learning algorithm, standard XGBoost, in this work, to take advantage of the ordering information contained in the hospital admission levels.

To utilise the ordered class values, the essential thought of this method is to transform a 𝑘-class ordinal problem into a 𝑘-1 binary classification problem. This is achieved by converting an ordinal attribute, denoted as 𝐴^∗^, with ordinal values 𝑉_1_, 𝑉_2_, 𝑉_3_, and so on up to 𝑉_𝑘_, into 𝑘 - 1 binary attributes. Specifically, for each of the first 𝑘-1 values of the original attribute, a binary attribute is created. Each of these binary attributes represents the test 𝐴^∗^ > 𝑉_𝑖_, where 𝑖 refers to the corresponding value of the original attribute. By adopting this method, we can effectively convert an ordinal regression problem into a binary classification problem, thereby enabling the application of various binary classification algorithms to the original ordinal problem.

The training process commences by generating novel datasets from the primary dataset, where a distinct dataset is generated for each of the 𝑘 - 1 newly formed binary class attributes. We take 𝑘 = 5 as an example here for illustration, i.e., hospital admissions are divided into five levels, where the hospitalisation increases as the value of levels increases. We then can convert it into 4 binary classification problems from the original dataset such that the binary target is 𝑖 if levels > 𝑖, so the classifier will predict *Pr (Level >* 𝑖*) where* 𝑖 *=1, 2, 3 and 4* Subsequently, the XGBoost algorithm is applied to generate a model for each of the newly created binary datasets separately. In order to predict the class value of an unseen instance, it is necessary to estimate the probabilities of the 𝑘 original ordinal classes, utilising the (𝑘 - 1) models developed in the previous step. The estimation of probabilities for the first and last ordinal class values is determined by a single classifier. The probability of the first ordinal value (*Level = 1*) is computed as 1 - *Pr (Level > 1)*. In the same manner, the probability of the last ordinal value (*Level = 5*) is determined by calculating the probability of *Pr (Level > 4)*. For class values that fall within the range between 1 and 5, the probability is given by a pair of classifiers. For example, the probability of Pr(*Level = 2*) is given by *Pr (Level > 1) - Pr (Level > 2)*.Generally, for any ordinal hospital admission level values 𝑉_𝑖_ the probability can be estimated as:

*Pr (*𝑉_1_) = 1- *Pr (Level >* 𝑉_1_)

*Pr (*𝑉_𝑖_) = *Pr (Level >* 𝑉_𝑖−1_) - *Pr (Level >* 𝑉_𝑖_ ), 1 < 𝑖 < 𝑘

*Pr (*𝑉_𝑘_) = 1- *Pr (Level >* 𝑉_𝑘−1_)

During the prediction phase, the (𝑘 - 1) classifiers are involved in calculating the probability of each of the k ordinal class values for an unknown instance by employing the aforementioned approach. The class value with the highest probability is assigned to the instance.

We applied the method described above to the standard XGBoost algorithm to construct a new model that uses ordering information, referred to as the XGBoost ordered model in the later section. The XGBoost ordered model is our primary forecast model and its performance is evaluated in comparison with other baseline models. In addition, the standard XGBoost, which treats each class attribute as a set of unordered values, was performed and compared its performance to the ordered XGBoost model. For the purpose of distinction, in subsequent sections, we refer to the standard XGBoost model as the XGBoost category model.

### Comparison null model

Ordered logistic regression (OLR) was employed as a baseline model to serve as a benchmark for evaluating the main model we explained above. The OLR model is an extension of logistic regression, which assumes proportional odds where the effect of the predictors is constant across all levels of the outcome variable [19]. It provides a simple but useful starting point for exploring the relationship between the predictors and the ordinal outcome. The predictors used in this model were limited to epidemiological predictors only (prior one- and two-week hospitalisation levels and prior one-week cases and deaths).

In addition to the OLR model, the null model in which the prediction of the target week is the same as the most recent available observation week, was used as a baseline model as well.

### Combinations of predictors

Hospital admission level is the ordinal outcome for each model. Three types of data were incorporated as predictors in the models: 1) epidemiological data, including ordinal hospitalisation levels, COVID-19 cases and deaths; 2) weather data, including temperature and total precipitation; 3) community mobility trends data in different types of locations. We only applied epidemiological data to the baseline models to provide baseline accuracy for model evaluation. For the XGBoost ordered and category models, we adapted four different combinations of predictors: 1) epidemiological data only; 2) epidemiological and weather data; 3) epidemiological and mobility data and 4) epidemiological, weather and mobility data. Detailed descriptions of data and predictors used for each model are summarised in Table 2.

**Table 2.**
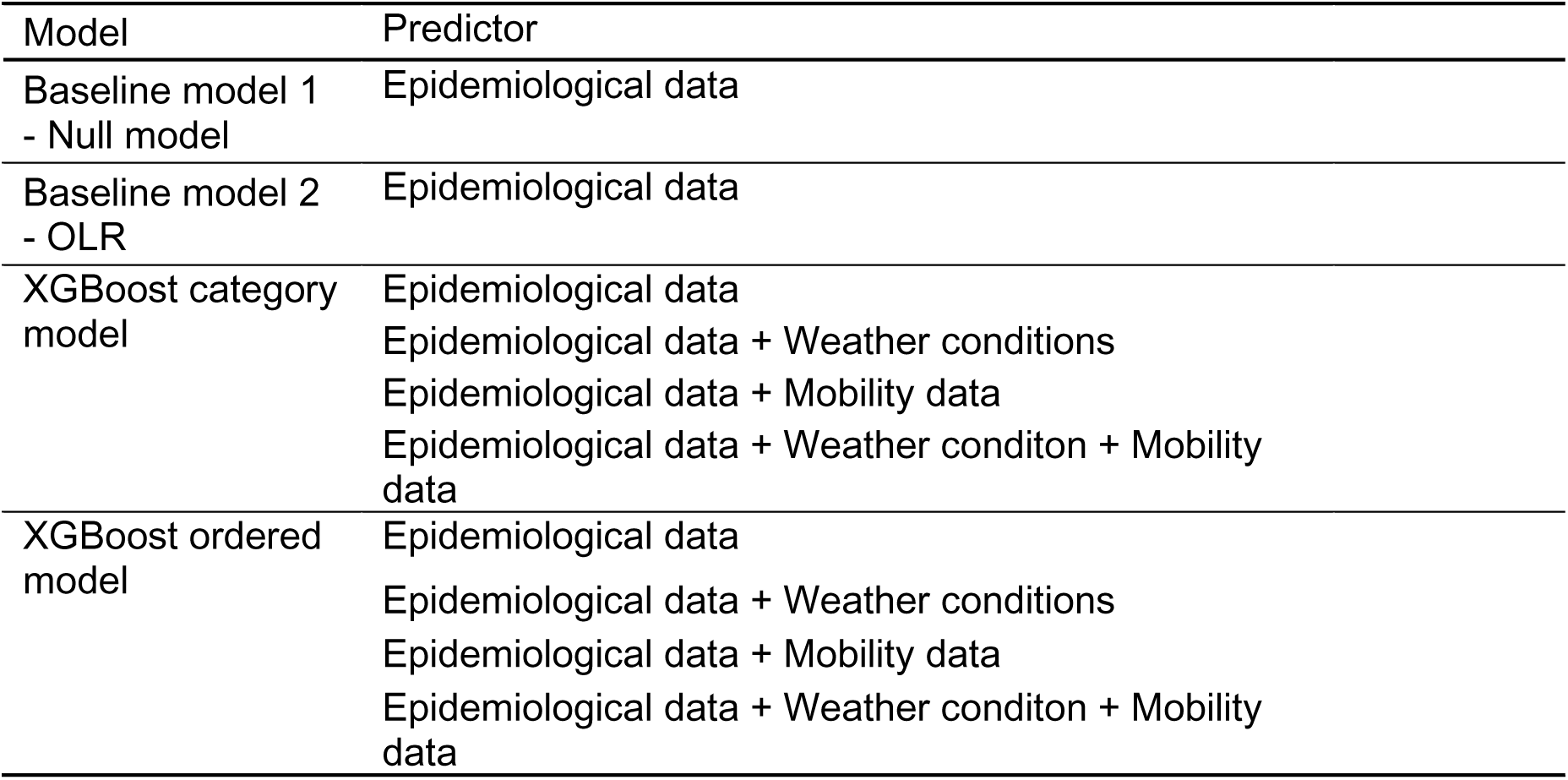
Summary of predictors incorporated by each model.

| Model | Predictor |
| --- | --- |
| Baseline model 1<br>- Null model | Epidemiological data |
| Baseline model 2<br>- OLR | Epidemiological data |
| XGBoost category<br>model | Epidemiological data<br>Epidemiological data + Weather conditions<br>Epidemiological data + Mobility data<br>Epidemiological data + Weather conditon + Mobility<br>data |
| XGBoost ordered<br>model | Epidemiological data<br>Epidemiological data + Weather conditions<br>Epidemiological data + Mobility data<br>Epidemiological data + Weather conditon + Mobility<br>data |

### Forecasting

The COVID-19 hospitalisation data can be characterised as a non-stationary time series, with a notable autocorrelation. Rather than employing a random data split for training and test sets, we made a deliberate choice. We designated the period from 9th March 2020 to 31st December 2021 as the training set, and the timeframe spanning from 1st January to 31st December 2022 as the test set. We also implemented the extending window approach, as previously detailed in our work [12] to facilitate multiple-step-ahead forecasts. In brief, we augmented the fitted period (i.e., the training set) with one new observation during each forecasting update. This methodology in defining training and test sets was consistently applied across all the models used in this work.

In terms of the hyperparameters for both the standard XGBoost model and XGboost ordered model, we maintained the same values that have been fine-tuned in our earlier work (Table S1) [12]. The analysis was conducted in R version 4.1.3.

### Evaluation metrics

We assessed the models based on two key metrics: macro-averaged mean absolute error (mMAE) and accuracy. Macro-averaged mean absolute error (mMAE) refers to the average of the mean absolute error (MAE) calculated for each class. More detailed explanations can be found in [12]. Accuracy is defined as the proportion of the number of weeks in which hospitalisation levels were correctly predicted to the total number of weeks within the test period.

## Results

Critical analysis of the COVID-19 pandemic in England reveals several significant trends related to the timing and magnitude of peaks in healthcare demand. These patterns are evident when examining both the raw hospitalisation data and an N-tile ordinal representation (Figure 1). Notably, during the dominance of the Alpha variant, hospitalisation reached its peak across all the NHS regions in January 2021. However, with the implementation of the third national lockdown and the rollout of vaccination programs and public health initiatives, hospitalisation rates gradually declined throughout the spring and summer months. The emergence of the Delta variants in mid-2021 marked a concerning shift as hospitalisations once again began to surge during the autumn and early winter of that year. Subsequently, the arrival of the Omicron variant in late 2021 ushered in yet another wave of increased hospitalisations, which persisted into the early months of 2022. Following this, the hospitalisation levels exhibited a fluctuating downtrend trajectory.

**Figure 1.**
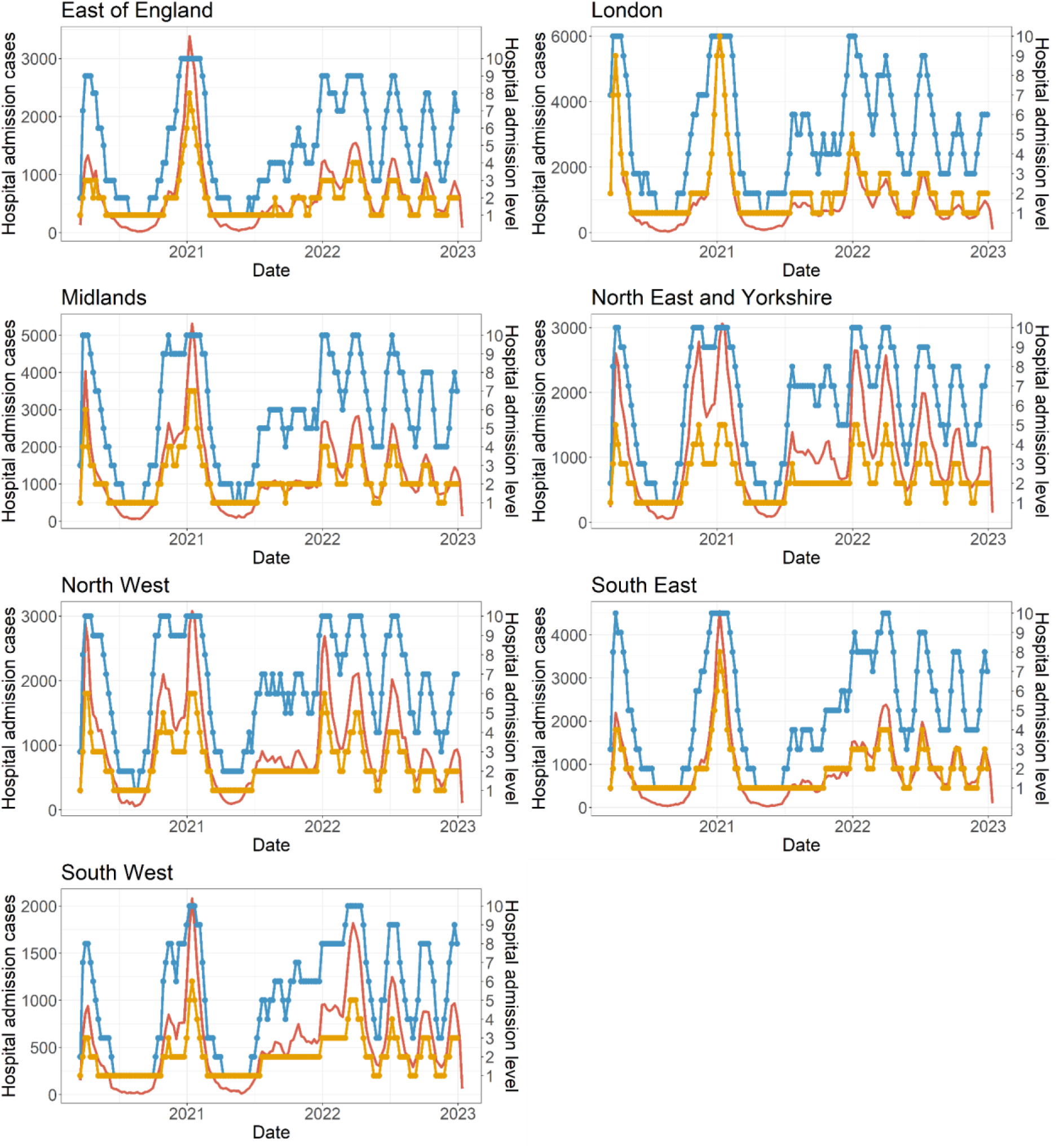
Epidemic curve of weekly hospitalisation by NHS regions in England from 19 March 2020 to 31 December 2022. The left y-axis represents the weekly count of new hospital admissions (red line), while the right y-axis depicts the weekly hospitalisation level determined using the N-tile method (blue line) and the uniform method (yellow line), with a total of 10 district levels.

Examining regional COVID-19 hospitalisation data from England provides a valuable opportunity to demonstrate the distinctions between the N-tile ordinal version of this data. Notably, the frequency distribution does not effectively differentiate between the ordinal levels as defined by the uniform method (S1 Figure). For instance, when using three levels, an overwhelming 90.02% of observations fall into level 1. With 5 levels, 94.81% of data points cluster within levels 1 and 2; while with 10 levels, a substantial 88% of the observations falls between 1 and 3. Consequently, the ordinal levels derived from the uniform approach appear to display a smoother and less fluctuating trend when compared to the actual numerical trend (Figure 1, S2 Figure). Therefore, we have opted to use the N-tile levels as our primary outcome and focus on assessing the predictive performance of models using the ordinal outcome established by the N-tile method. We will revisit the potential impact of this choice in a subsequent sensitivity analysis.

Our initial evaluation of model performance exclusively considered epidemiological data, which may be the only option for many populations during future similar periods. Surprisingly, we consistently achieved more accurate forecasts by the XGboost ordered model when compared to baseline models for 1- to 4-week ahead predictions even without incorporating additional potential features such as weather and mobility in the model (S3 Figure).

Subsequently, we introduced weather and mobility data as predictors into both the XGBoost ordered and category models evaluating model performance by macro-averaged Mean Absolute Error (mMAE). The mMAE was computed as the average value across seven NHS regions. Incorporating weather data alongside epidemiological data indeed improved the performance of our forecasting models when compared to baseline models (Figure 2). However, when comparing XGBoost models featuring both weather and epidemiological data with those utilising only epidemiological data, there was no improvement in accuracy (S4 Figure).

**Figure 2.**
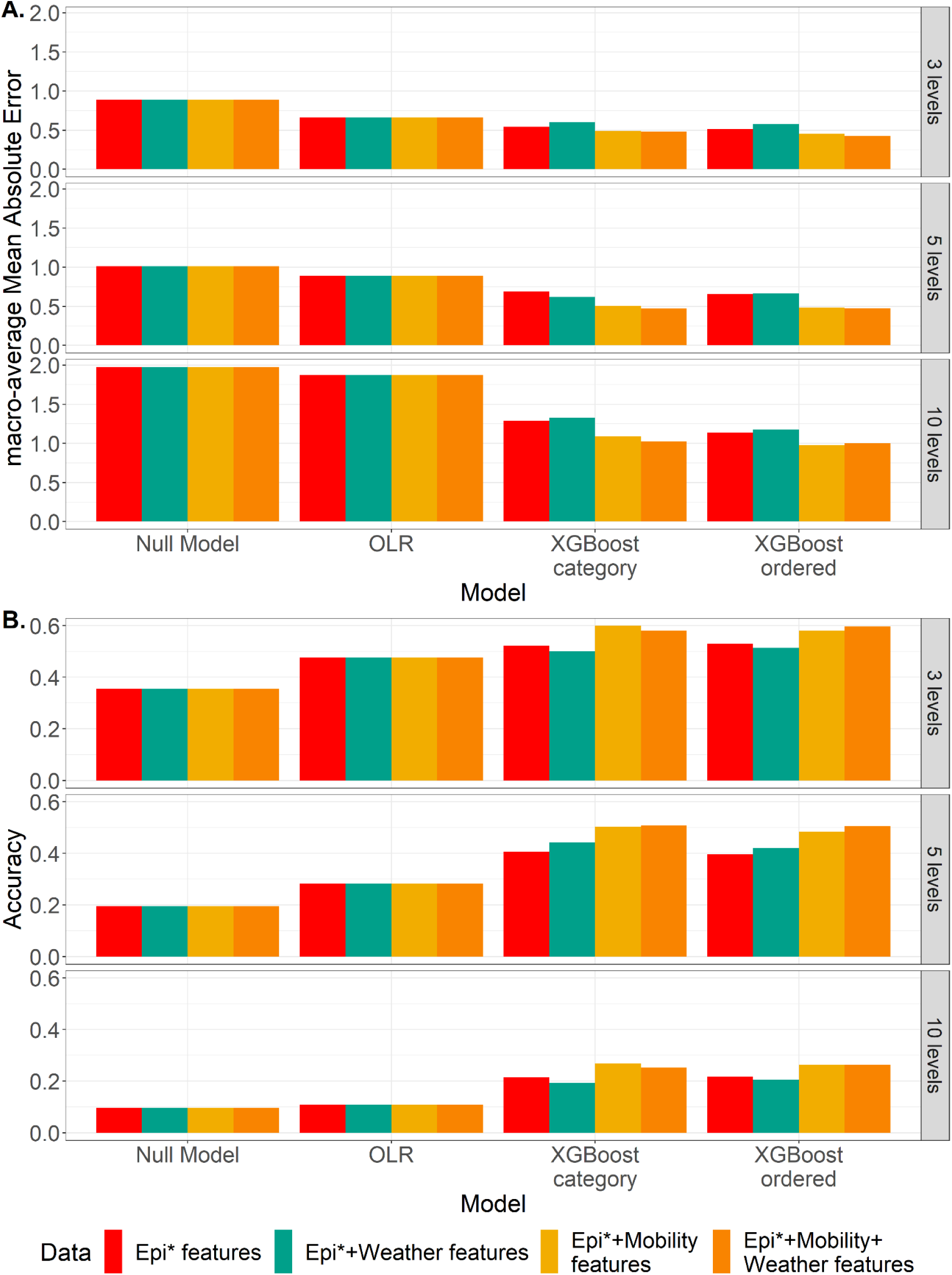
Accuracy metrics comparison of 4-week ahead forecasts between models trained with different combinations of predictors. A. Average macro-averaged Mean Absolute Error (mMAE) over all seven NHS regions. **B.** Average accuracy over all seven NHS regions. *Epidemiological

In contrast, our findings suggest that the mobility data significantly enhances forecast accuracy compared to relying solely on epidemiological data alone (Figure 2). Notably, the XGBoost model that exclusively incorporates epidemiological data consistently performed notably worse than those integrating mobility data (S5 Figure). This underscores the importance of mobility data as a crucial factor in COVID-19 hospitalisation forecasting models.

Finally, we incorporated both mobility and weather data into the models. Figure 3 illustrates the average mMAE for each of the four combinations of predictors in the XGBoost models across various prediction horizons (1- to 4-week ahead). Within each prediction horizon, mMAE increases as the prediction horizon increases, highlighting the increased challenge of making predictions further ahead in time. Nevertheless, across all prediction horizons, models that incorporate mobility data consistently demonstrate superior performance. Particularly noteworthy is the reduction in mMAE for 3- and 4-week ahead prediction when both weather and mobility are used as predictors.

**Figure 3.**
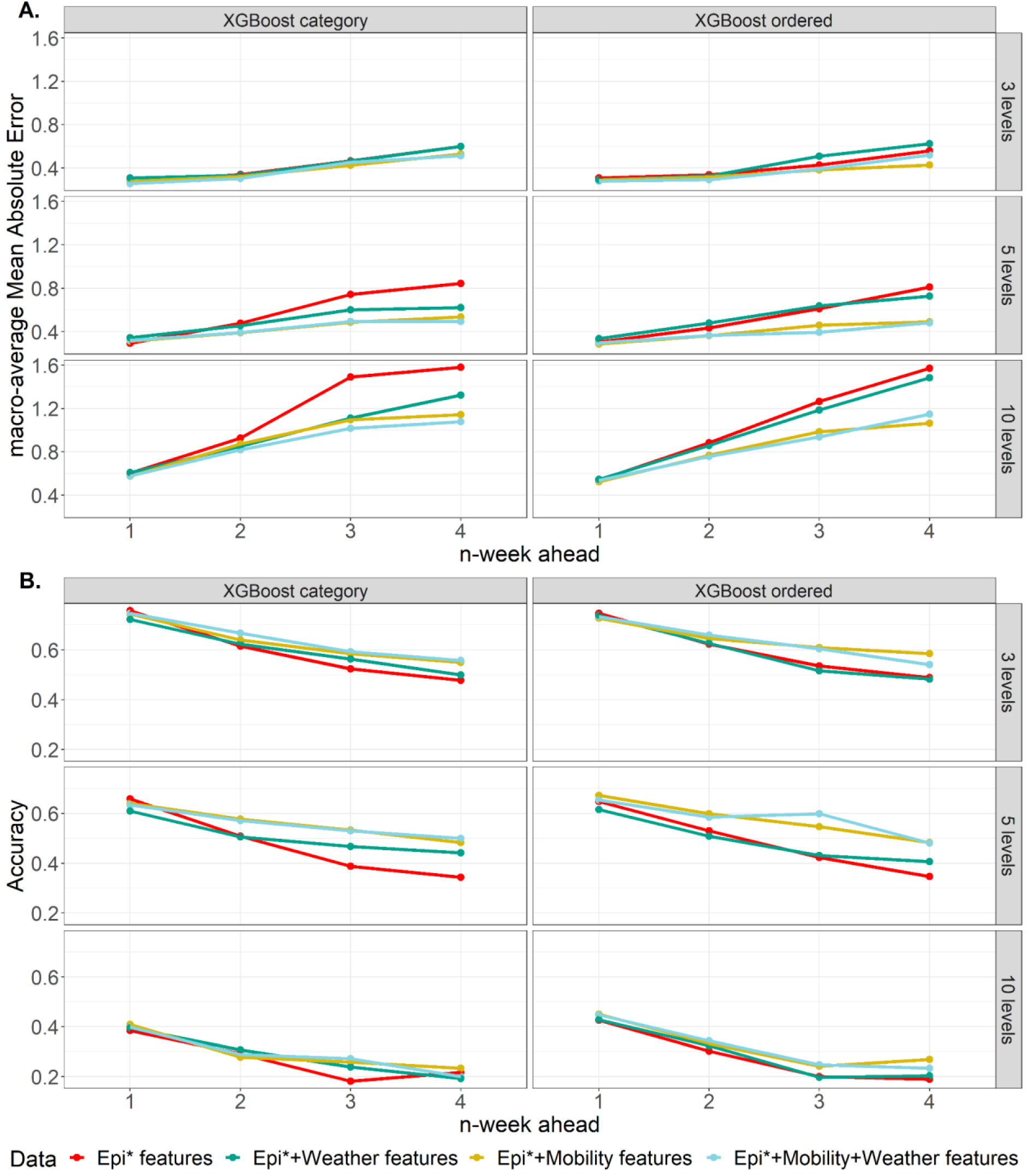
Evaluating XGBoost model performance with various feature sets, using macro-averaged Mean Absolute Error (mMAE) and accuracy. The overall mMAE was computed by averaging across the seven NHS regions for each prediction horizon spanning from 1- to 4-week ahead). **A.** mMAE of both the XGBoost category model and the XGBoost ordered model trained with the following sets of features 1) epidemiological data (red line); 2) epidemiological and weather data (green line); 3) epidemiological and mobility data (yellow line); and 4) epidemiological, mobility and weather features (blue line). **B.** Accuracy results for the XGBoost category model and the XGBoost ordered model, which was trained using the same feature sets as in Panel A with 1) epidemiological data (red line); 2) epidemiological and weather data (green line); 3) epidemiological and mobility data (yellow line); and 4) epidemiological, mobility and weather features (blue line). *Epidemiological

We also investigated the relative merits of different numbers of ordinal levels and Table 2 summarises the results for the 3-, 5-, and 10-bin levels based on the N-tile bins. It becomes evident that the XGBoost ordered model consistently outperforms the baseline models in terms of accuracy for both 3 and 5 bins defined by the N-tile method. However, it is interesting to note that the XGBoost ordered model does not always exhibit a significantly superior performance. For the 10-bin scenario, we observe more substantial differences in accuracy (Table 3, Figure 3), in particular when comparing it to the XGBoost category model under the N-tile method. In this context, the divergence in performance between the XGBoost ordered model and the baseline models becomes more significant. Moreover, the XGBoost surpasses the XGBoost category model across all four predictor combinations and prediction horizons. This substantial improvement underscores the increasing relevance of ordering information as the number of classes increases (Figure 4).

**Figure 4.**
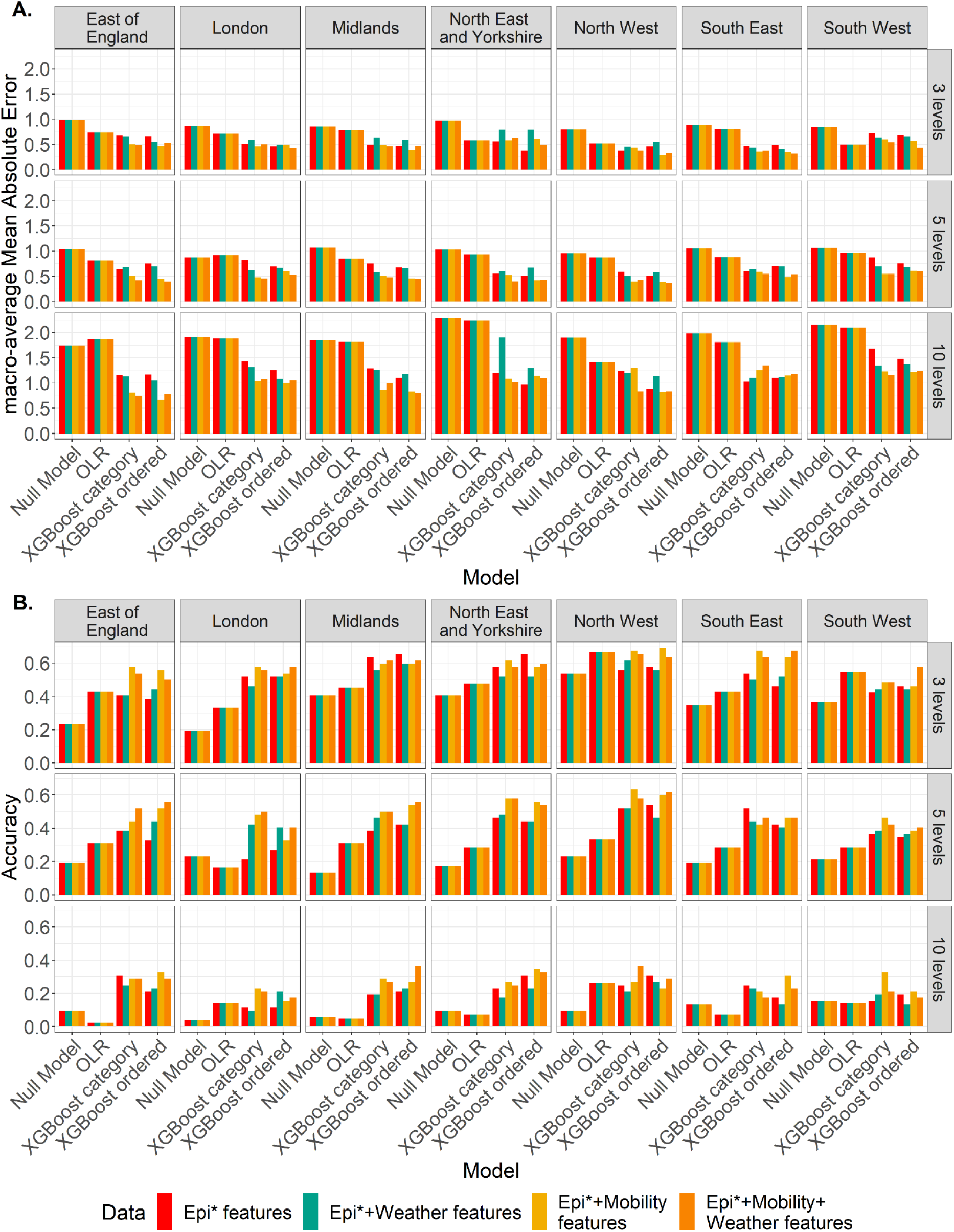
Comparison of model performance by NHS regions for XGBoost models and baseline models trained with different combinations of predictors when the prediction horizon is 4 week-ahead for the test period 2022. The levels are defined by the N-tile method. **A.** mMAEs of forecasts for hospitalisation levels. **B.** Accuracy of forecasts for hospitalisation levels. *Epidemiological

**Table 3.** Predictive performance of models when the N-tile method is used to define the ordinal level of hospital admissions.

| n-week<br>ahead |  | Bins = 3 |  | Bins = 5 |  | Bins = 10 |  |
| --- | --- | --- | --- | --- | --- | --- | --- |
|  |  | mMA<br>E | Accuracy (%) | mMA<br>E | Accuracy (%) | mMA<br>E | Accuracy (%) |
| 1-week<br>ahead | Null model | 0.294 | 76.92% | 0.290 | 66.48% | 0.604 | 39.29% |
|  | OLR | 0.302 | 73.47% | 0.317 | 60.88% | 0.498 | 44.56% |
|  | XGBoost<br>category | 0.298 | 72.80% | 0.305 | 64.56% | 0.605 | 37.91% |
|  | XGBoost<br>ordered | 0.293 | 71.43% | 0.279 | 67.03% | 0.494 | 45.60% |
| 2-week<br>ahead | Null model | 0.536 | 56.59% | 0.540 | 43.41% | 1.088 | 23.35% |
|  | OLR | 0.503 | 55.78% | 0.424 | 47.28% | 0.960 | 24.49% |
|  | XGBoost<br>category | 0.315 | 65.38% | 0.376 | 58.79% | 0.811 | 31.87% |
|  | XGBoost<br>ordered | 0.300 | 65.93% | 0.340 | 61.26% | 0.668 | 35.44% |
| 3-week<br>ahead | Null model | 0.717 | 42.03% | 0.793 | 28.57% | 1.593 | 13.46% |
|  | OLR | 0.540 | 54.08% | 0.624 | 38.78% | 1.366 | 18.37% |
|  | XGBoost<br>category | 0.349 | 61.81% | 0.473 | 55.77% | 0.971 | 29.40% |
|  | XGBoost<br>ordered | 0.346 | 64.56% | 0.388 | 60.44% | 0.832 | 28.85% |
| 4-week<br>ahead | Null model | 0.888 | 35.44% | 1.011 | 19.51% | 1.974 | 9.62% |
|  | OLR | 0.663 | 47.62% | 0.891 | 28.23% | 1.874 | 10.88% |
|  | XGBoost<br>category | 0.483 | 57.97% | 0.470 | 50.82% | 1.024 | 25.27% |
|  | XGBoost<br>ordered | 0.427 | 59.62% | 0.475 | 50.55% | 1.002 | 26.37% |

To assess the significance of predictors, we used “the Gain”, a metric that quantifies the relative contribution of each feature to the XGBoost ordered model. The five most influential predictors are as follows: hospitalisation levels from previous weeks, the number of COVID-19 cases, temperature, mobility changes in retail and recreation places, and the number of COVID-19-associated deaths (Figure S6). These key predictors underscored the importance of epidemiological data and how the inclusion of weather and mobility data further enhances the model’s accuracy. Conversely, total precipitation and other mobility trends exhibit lower importance in the model.

Our findings are corroborated when we compare results across individual regions (Figure 5). Using the N-tile method to define hospitalisation levels, we predicted these levels 1- to 4- week ahead throughout the entire test period. Notably, the prediction errors (mMAEs) are consistently lower in models that t feature mobility data and even lower when both mobility and weather data are included, as compared to using only epidemiological or weather alone for all regions. As the ordinal class value increases, the discrepancy in prediction errors becomes more pronounced. For instance, in the East of England, when the class number is set to 10, the mMAE of the 2-week ahead forecast only uses epidemiological data is about double that of a 4-week ahead forecast incorporating both weather and mobility data. When extending the prediction horizon to 4-week ahead, the difference in mMAE increases threefold.

**Figure 5.**
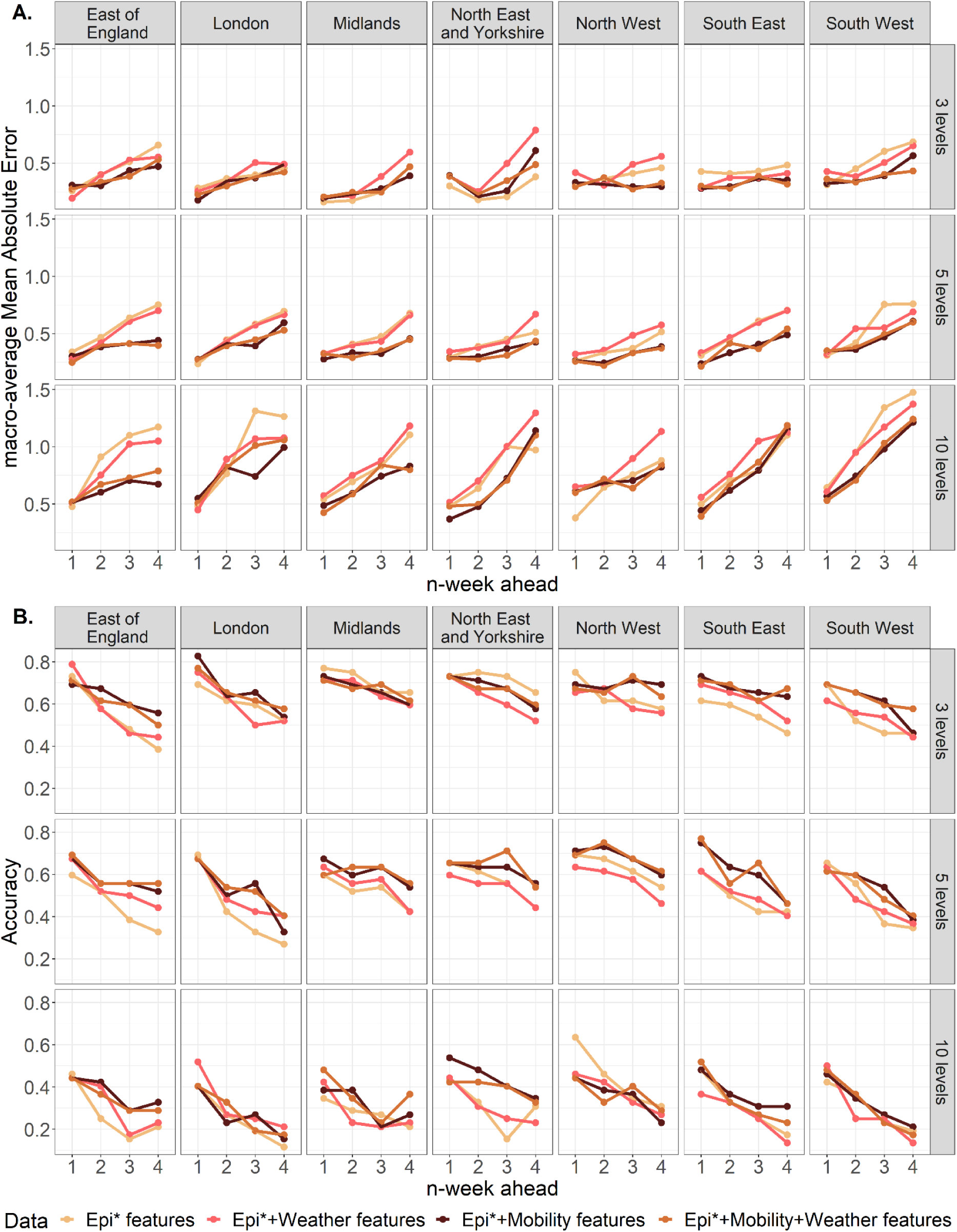
Comparison of NHS-region-specific performance for the XGBoost ordered model trained with 1) epidemiological features (beige line); 2) epidemiological and weather features (pink line); 3) epidemiological and mobility features (brown line); and 4) epidemiological, weather and mobility features (yellow line). The prediction horizon is from 1 to 4 week-ahead for the test period 2022. The number of total levels orders the rows from 3 (top) to 10 (bottom) levels, and levels are defined by the N-tile method. Each column represents one NHS region. **A.** mMAEs of forecasts for hospitalisation levels. **B.** Accuracy of forecasts for hospitalisation levels. *Epidemiological

In Figure 6, we conducted a comparative analysis between the predicted values generated by the XGBoost ordered models and the time series data representing actual hospitalisation values. Across all NHS regions, all four predictor combinations effectively capture the overall hospitalisation trends and successfully identify the peak admission levels for 1- and 2-week forecasts. Particularly noteworthy is the superior accuracy of models incorporating mobility data and models combining both mobility and weather data. However, when looking at 3- and 4-week ahead forecasts, the model relying solely on epidemiological data exhibits noticeable deviations from the actual levels, primarily evident in a lag in predicting peak admissions. For these longer-term forecasts, the models integrating mobility data, and especially those incorporating both mobility and weather data, do not entirely capture the fluctuations observed at the beginning of 2020. Nonetheless, their predictions for up to 4-weeks ahead remain the closest approximation to the actual observed peak, emphasizing their predictive strength even in the face of more extended forecasting horizons.

**Figure 6.**
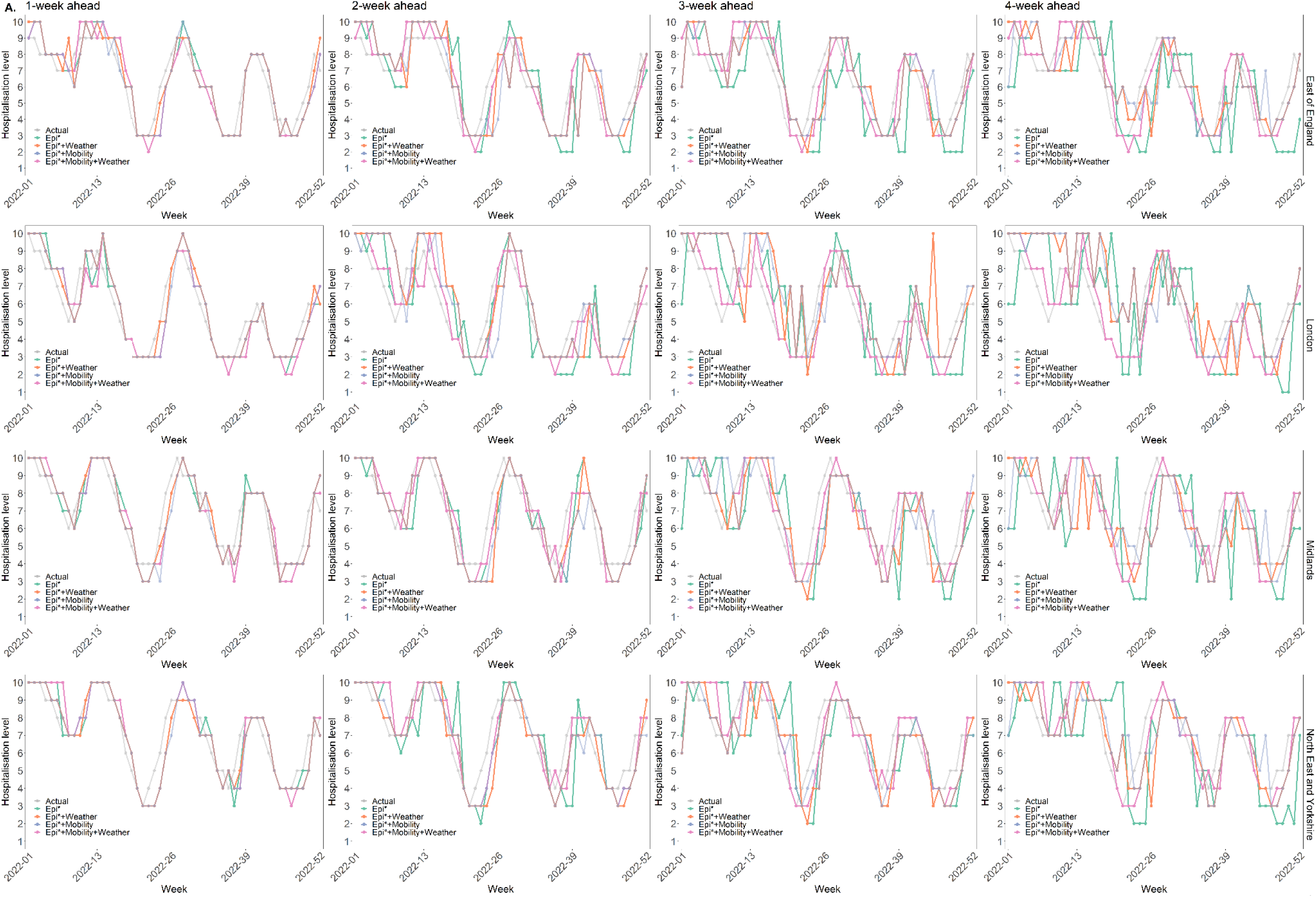
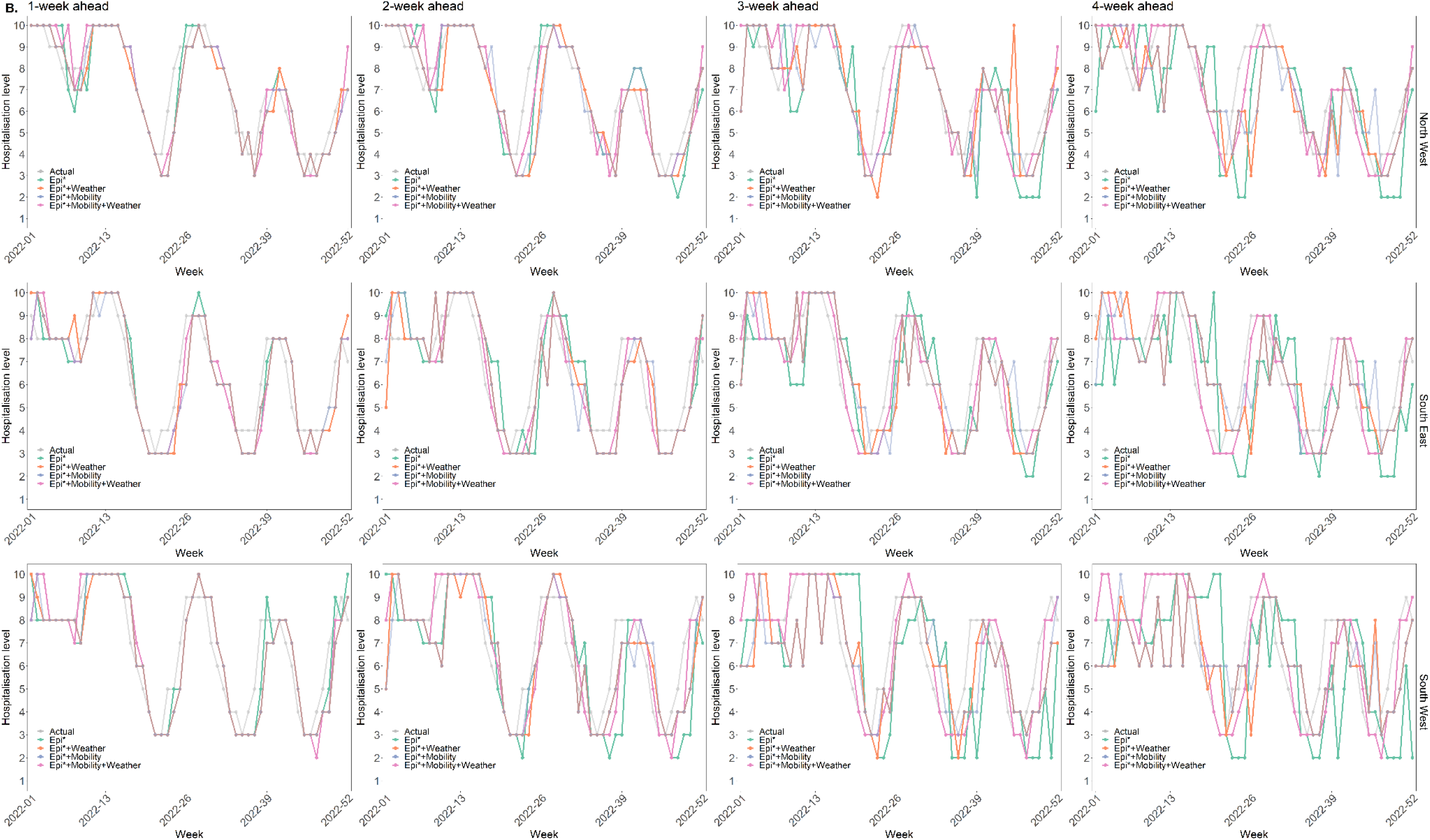
Comparison of actual and predicted hospitalisation levels by NHS regions. The predicted hospitalisation levels are predicted by the XGBoost ordered model with 10 levels defined by the N-tile method, and the time ranges from week 1 to week 52 in 2022. Each row represents one NHS region. Columns are prediction horizons ordered from 1- (left) to 4-week (right) ahead. **A.** East of England, London, Midlands and North East and Yorkshire; **B.** North West, South East and South West.

## Discussion

This study aims to enhance the accuracy of forecasting regional hospitalisation levels in England, which are treated as ordinal outcomes, by integrating various data sources. We evaluated the efficacy of different bin partitioning methods in leveraging the inherent ordering information Notably, the N-tile method outperforms the uniform method in this regard. Our study covered seven NHS regions and assesses the performance of short-term forecasts in three distinct scenarios: 1) forecasts solely reliant on epidemiological data, including previous hospitalisation figures, case counts, and mortality statistics; 2) forecasts that incorporate mobility data and weather conditions as supplementary predictors, considered separately from epidemiological data; 3) forecasts that incorporate both mobility data and weather conditions as additional predictors alongside epidemiological data.

Our findings reveal that the XGboost ordered model, based solely on epidemiological data, achieves superior accuracy compared to baseline models. This outcome is particularly beneficial for countries with limited access to diverse data sources beyond epidemiological data. Furthermore, we provide evidence that the inclusion of aggregated mobility data significantly enhances prediction accuracy, extending up to a 4-week horizon when compared to models relying solely on epidemiological data. However, our analysis indicates that the contribution of weather conditions to prediction accuracy is minimal.

A key insight from our study is the strong predictive power of mobility data in relation to COVID-19 transmission compared to weather conditions. Our findings are consistent with prior research that underscored the pivotal role of mobility as a primary predictor of COVID-19 transmission dynamics [20–23]. Several other studies have also explored the association between mobility and COVID-19 transmission but have arrived at varying conclusions. For instance, a study conducted by[24] found mobility to be a significant predictor of COVID-19 cases in specific regions, while observing limited impact in others. Meanwhile, [25] demonstrated a strong correlation between mobility and COVID-19 transmission across diverse geographical areas. These disparities in findings may be attributed to differences in study design, regional or geographical contexts, and specific modelling approaches employed. Differences in data collection methods and time periods considered in each study may also contribute to the observed discrepancies. In our analysis, we believe that the extensive and diverse dataset we used, allowed us to discern mobility as the primary predictor. Additionally, the incorporation of local factors and spatiotemporal variations in mobility patterns might have contributed to our more accurate predictions.

Mobility data reflects the contact behaviour changes occurring within a population in response to COVID-19, while weather factors may interact with these changes and contribute to the spread of COVID-19, potentially leading to increased hospitalisation. For example, during mild weather conditions, individuals tend to spend more time outdoors in parks and engage in socialising activities involving close contact without adhering to safety measures such as wearing masks and physical distancing.

Nonetheless, our study has its limitations. First, we assumed that aggregated mobility data captures well the human social mixing patterns in the population. The robustness of our model forecasts relies heavily on the accessibility and representativeness of this aggregate mobility data. It is important to note that Google ceased updating their mobility data from 15 October 2022 onwards, which may limit the generalisability of our results in the future due to the reduced availability of mobility datasets. Moreover, in resource-constrained settings with limited smartphone usage, the aggregated mobility data may not fully characterise the movement patterns of the entire population.

Furthermore, the lack of continuous and long-term availability of mobility or weather data in certain resource-constrained regions or countries is another constraint. Consequently, the generalisability of our findings may be impacted, as the reduced availability of mobility and weather datasets could hinder the accurate assessment and prediction of the transmission dynamics of COVID-19 in those areas. For these reasons, we also described here the performance of the XGBoost model of ordinal data using only intrinsic variables in the time series.

We were unable to obtain sufficiently accurate datasets to consider the incorporation of non-pharmaceutical interventions (NPIs) and vaccination as additional features in our predictive model at the regional level. Specifically, the absence of suitable weekly vaccination data by NHS regions hindered our ability to assess the potential influence of vaccination coverage rates on the accuracy of our predictions. Furthermore, we did not include factors such as the effectiveness of NPIs and behavioural patterns, such as intentions of wearing face masks and following social distancing, as predictors in our model. However, it is widely acknowledged that vaccination and non-pharmaceutical interventions have played pivotal roles in mitigating the spread of COVID-19 and reducing infection and hospitalisation rates in various regions[26,27]. The rollout of vaccination campaigns and the implementation of NPIs, including lockdowns, travel restrictions, and mask mandates, have demonstrated their efficacy in curbing transmission and alleviating the strain on healthcare systems[28,29]. We propose that the framework we have outlined here could be effectively employed when such data become available, allowing for the assessment of the potential value of these variables in predicting critical hospitalisation patterns.

Considering the significant impact of vaccination and NPIs on disease transmission dynamics, we acknowledge the critical importance of integrating these factors in future modelling efforts. The inclusion of vaccination coverage rates, NPI implementation timelines, and compliance levels with behavioural interventions would enrich our predictive model and enhance its accuracy in forecasting COVID-19 trends at the regional levels. These enhancements could facilitate more precise and timely public health decision-making and resource allocation during pandemic management. To advance this effort, it is imperative to have access to available and reliable datasets encompassing vaccination coverage and NPI implementation across NHS regions. Robust data collection and reporting mechanisms are essential for researchers and policymakers to gain a comprehensive understanding of the interplay between vaccination efforts, NPI adoption, and disease transmission dynamics[30,31]. Collaborative initiatives among healthcare agencies, governmental bodies, and research institutions are also important for establishing standardised data collection protocols and enabling the timely sharing of accurate information.

In conclusion, our findings underscore the value of incorporating weather and mobility data to explore the ordering information of ordinal hospitalisation levels, thereby enhancing the precision of hospital admission level predictions over a 4-week ahead time horizon. This extension provides policymakers with additional time to plan and allocate hospital resources effectively.

## Supporting information

Main Tables 1-2

## Supporting information

S1 Table. Hyperparameter values used by the XGBoost models.

(XLSX)

S2 Table. Predictive performance of models when the ordinal level of hospital admissions is defined by the uniform method.

(XLSX)

**S1 Figure.**
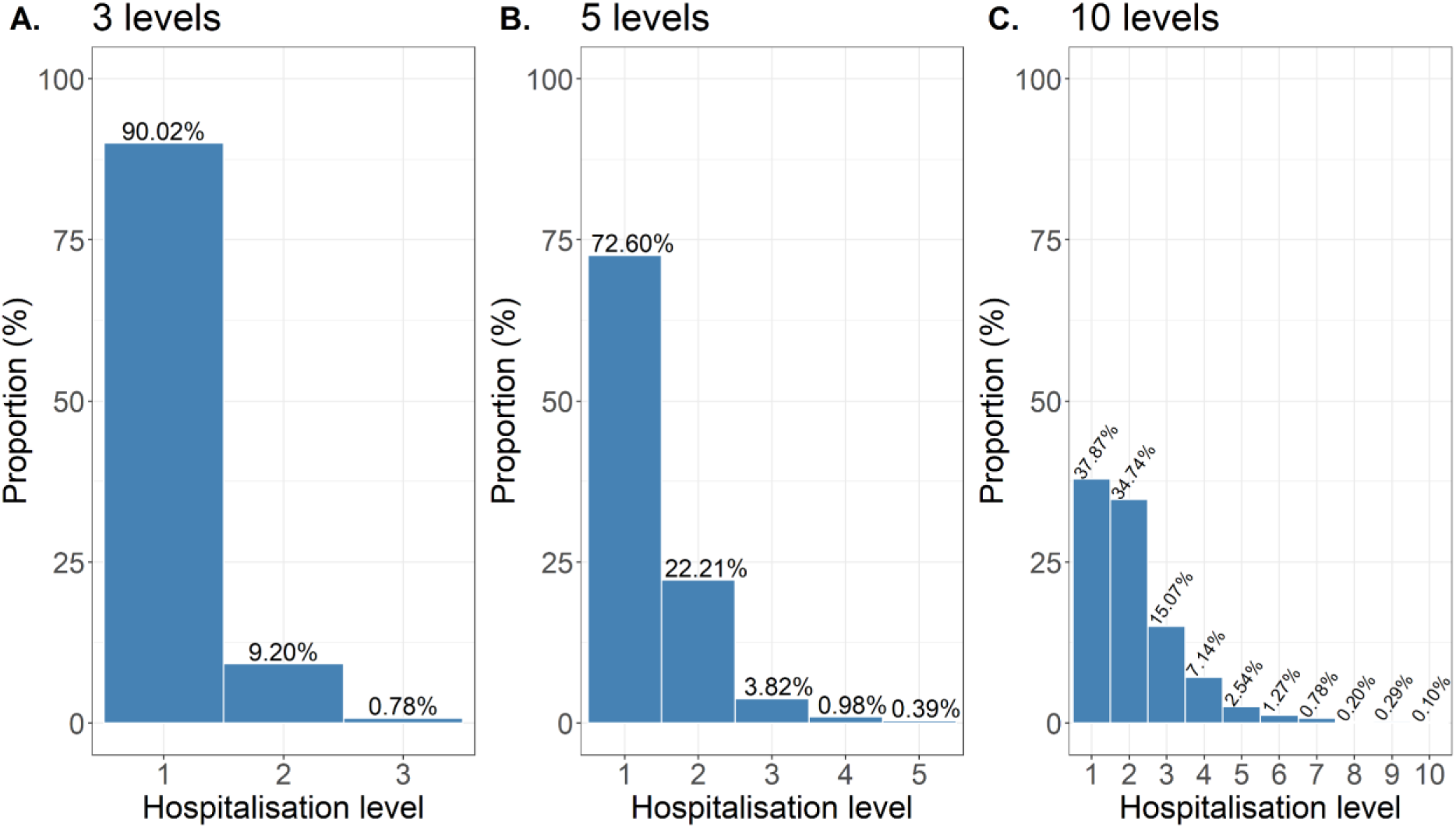
Distribution of ordinal levels for weekly hospitalisations for NHS regions discretised by the uniform method.

**A.** The distribution of hospital admission that discretised into three levels. **B.** The distribution of hospital admission that discretised into five levels. **C.** The distribution of hospital admission that discretised into ten levels.

**S2 Figure.**
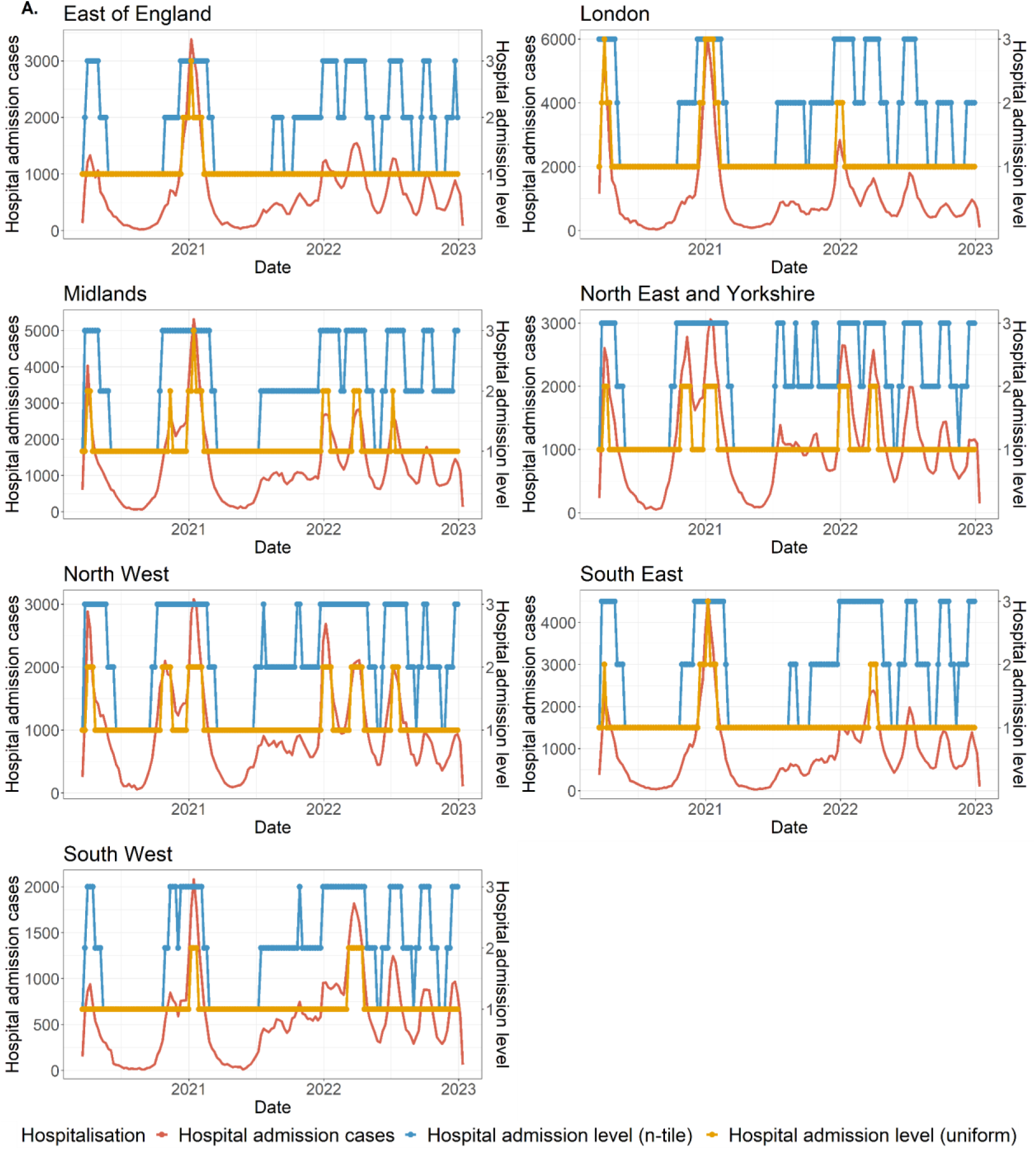
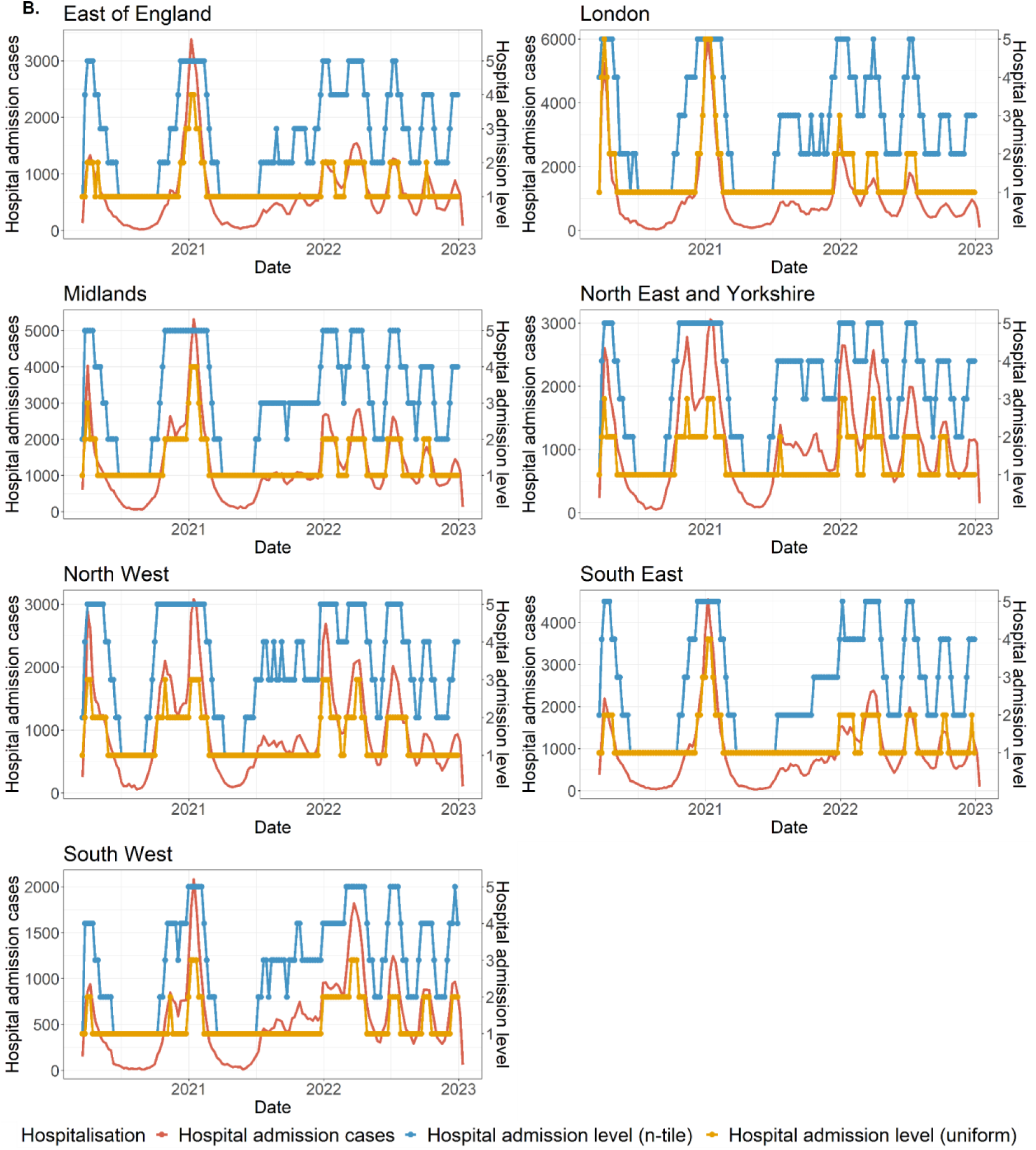
Epidemic curve of weekly hospitalisation by NHS regions in England from 19 March 2020 to 31 December 2022.

The left y-axis is the numerical number of weekly new hospital admissions (red line), while the right y-axis is the ordinal weekly hospitalisation level defined by the N-tile method (blue line) and the uniform method (yellow line). **A.** The number of levels equals three. **B.** The number of levels equals five.

**S3 Figure.**
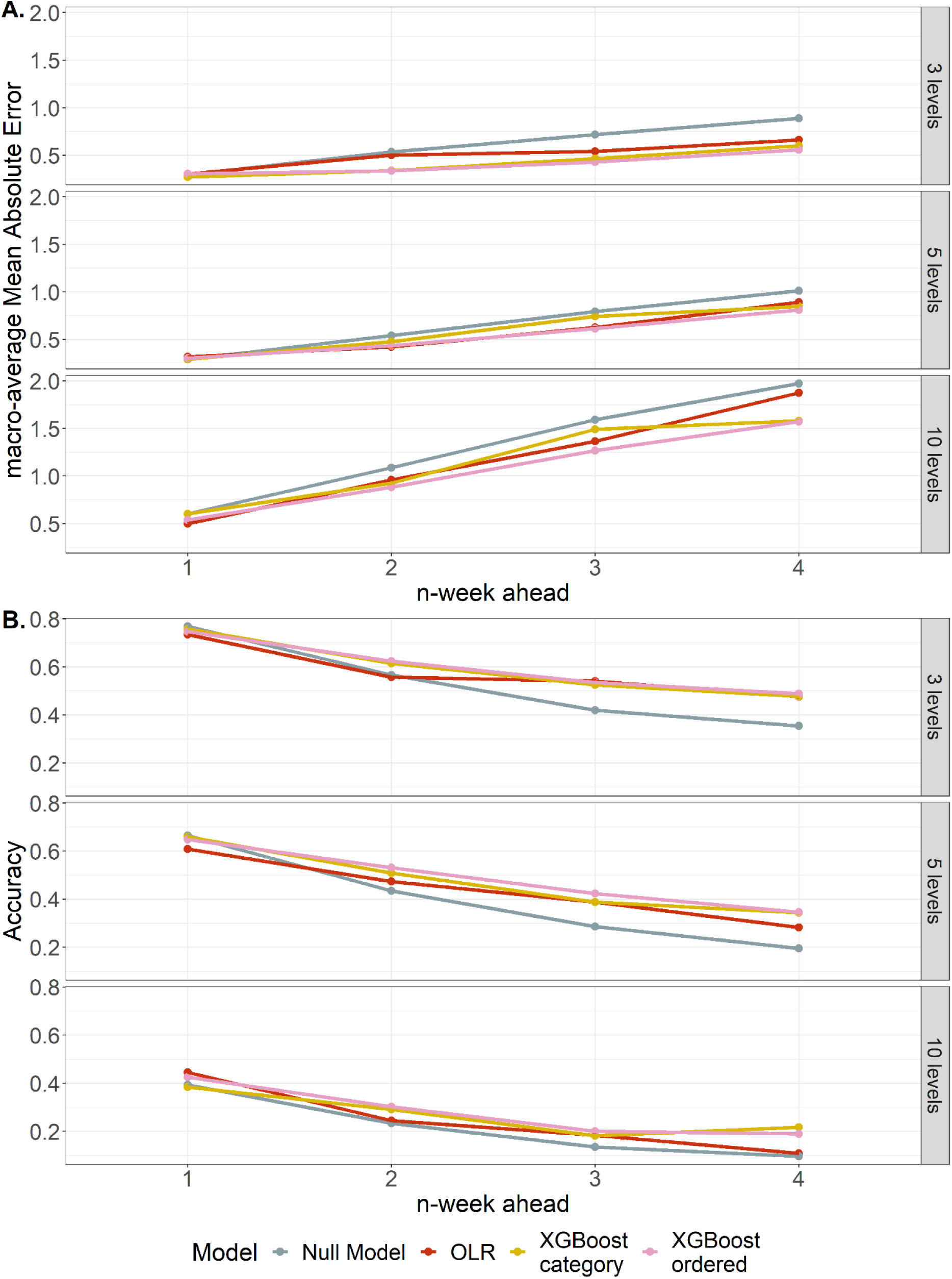
Model performance of predicting N-tile hospitalisation levels with only epidemiological predictors.

**A.** mMAEs of the null model (grey line), ordered logistic regression model (red line), the XGBoost category model (yellow line) and the XGBoost model (pink line). **B.** Accuracy of the null model (grey line), ordered logistic regression model (red line), the XGBoost category model (yellow line) and the XGBoost model (pink line).

**S4 Figure.**
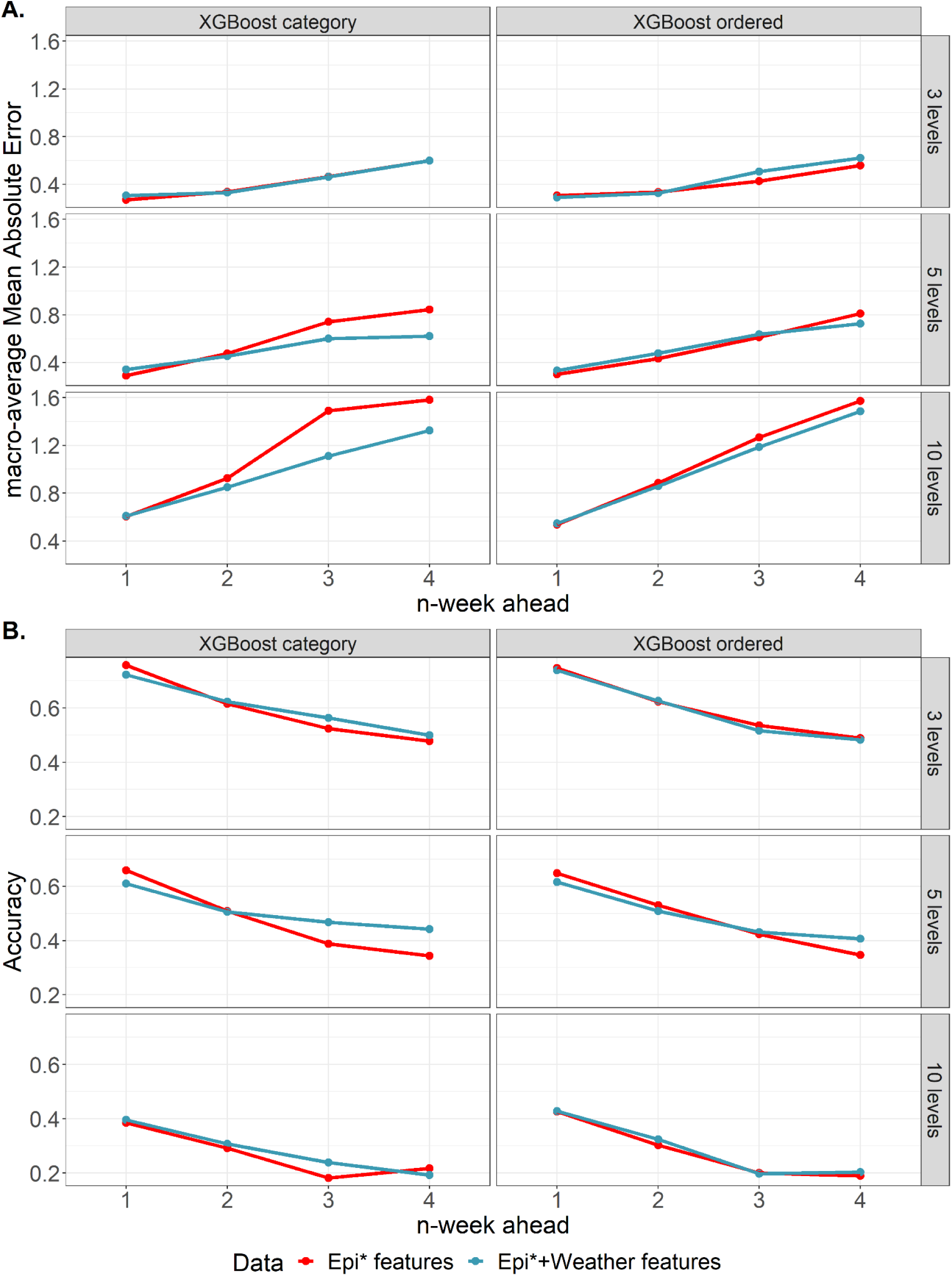
Performance of the XGBoost models incorporated with epidemiological and weather data, measured by macro-averaged Mean Absolute Error (mMAE) and accuracy.

The overall mMAE and accuracy were averaged over seven NHS regions for each prediction horizon (1- to 4-week ahead). **A.** mMAEs of the XGBoost models trained with epidemiological and weather features (blue line) and epidemiological features (red line). **B.** Accuracy of the XGBoost models trained with epidemiological and weather features (blue line) and epidemiological features (red line).

*Epidemiological

**S5 Figure.**
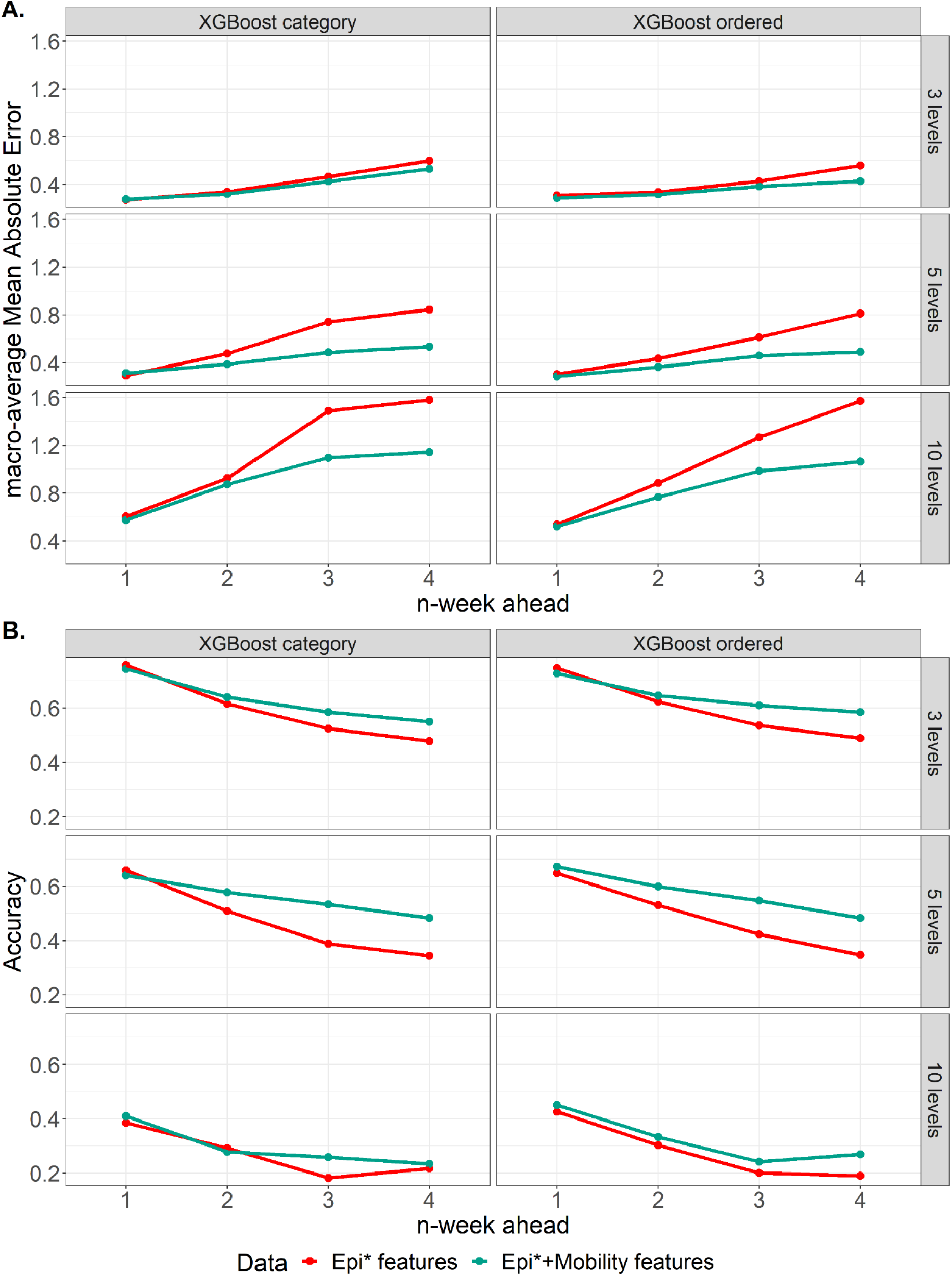
Performance of the XGBoost models incorporated with epidemiological and mobility data, measured by macro-averaged Mean Absolute Error (mMAE) and accuracy.

The overall mMAE and accuracy were averaged over seven NHS regions for each prediction horizon (1- to 4-week ahead). **A.** mMAEs of the XGBoost models trained with epidemiological and mobility features (green line) and epidemiological features (red line). **B.** Accuracy of the XGBoost models trained with epidemiological and mobility features (green line) and epidemiological features (red line).

*Epidemiological

**S6 Figure.**
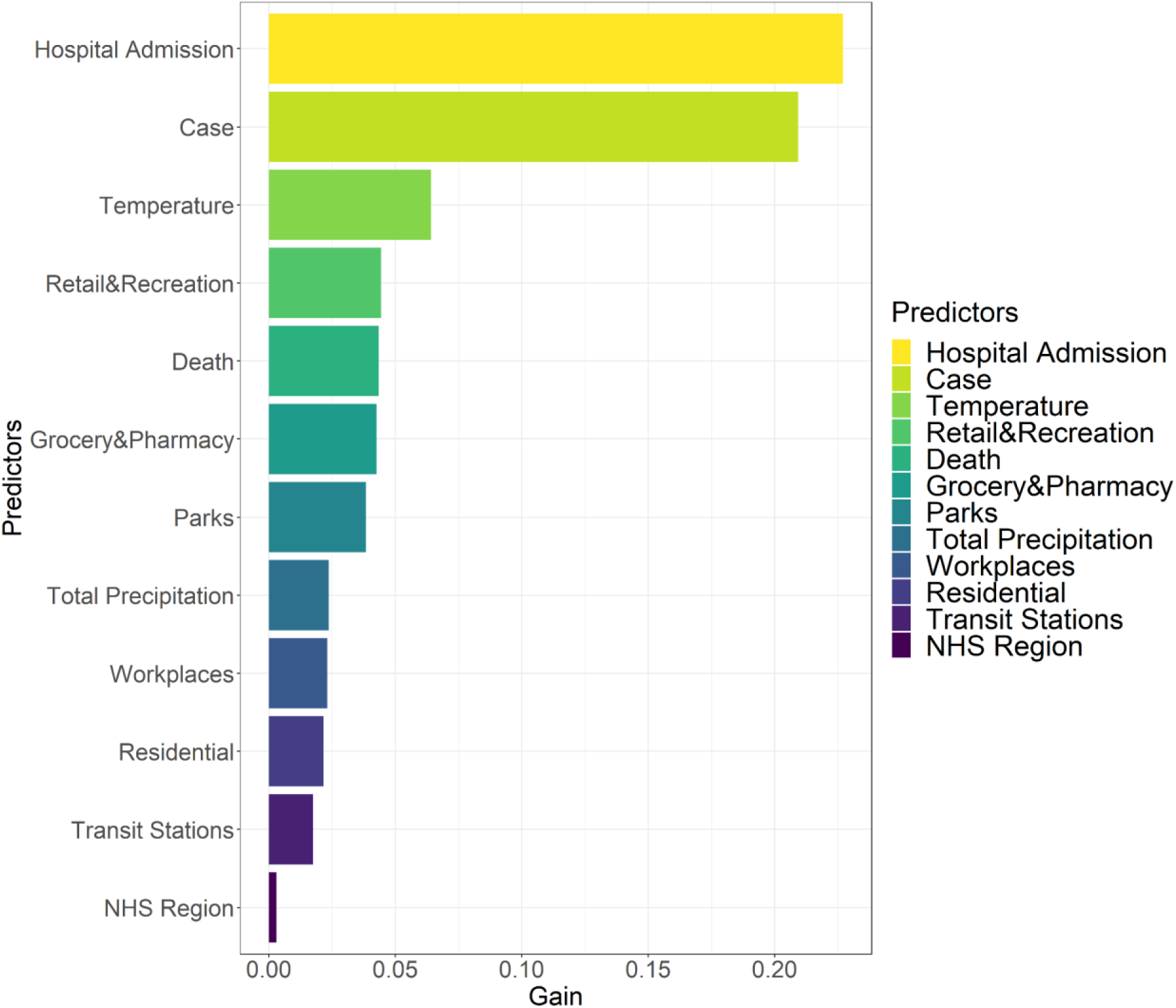
The relative importance (Gain) of predictors used by the XGBoost ordered model.

## Data availability

The source code and data used to produce the results and analyses presented in this manuscript and supporting figures and tables are available on a Git repository: https://github.com/VVVVivi/COVID_hosp_forecasting.git. Supporting figures and tables are provided in the same GitHub repository as extended data.

## Acknowledgements

The authors acknowledge funding from the MRC Centre for Global Infectious Disease Analysis (MR/R015600/1), jointly funded by the UK Medical Research Council (MRC) and the UK Foreign, Commonwealth & Development Office (FCDO), under the MRC/FCDO Concordat agreement, and also part of the EDCTP2 programme supported by the European Union. S.R. acknowledges the support from Wellcome Trust Investigator Award (UK, 200861/Z/16/Z). KOK acknowledges funding from HMRF (INF-CUHK-1). RL acknowledges funding from Nanjing Medical University Talents Start-up Grants (NMUR20220001).

## Author Contribution

**Conceptualization:** Haowei Wang, Steven Riley

**Data curation:** Haowei Wang

**Formal analysis:** Haowei Wang

**Methodology:** Haowei Wang

**Software:** Haowei Wang

**Supervision:** Steven Riley

**Validation:** Haowei Wang

**Visualization:** Haowei Wang

**Writing – original draft:** Haowei Wang

**Writing – review & editing:** Haowei Wang, Steven Riley, Kin On Kwok, Ruiyun Li

