## Supplementary figures and images for "Forecasting regional-level COVID-19 hospitalisation in England as an ordinal variable using the machine learning method"

### S1 Figure.png

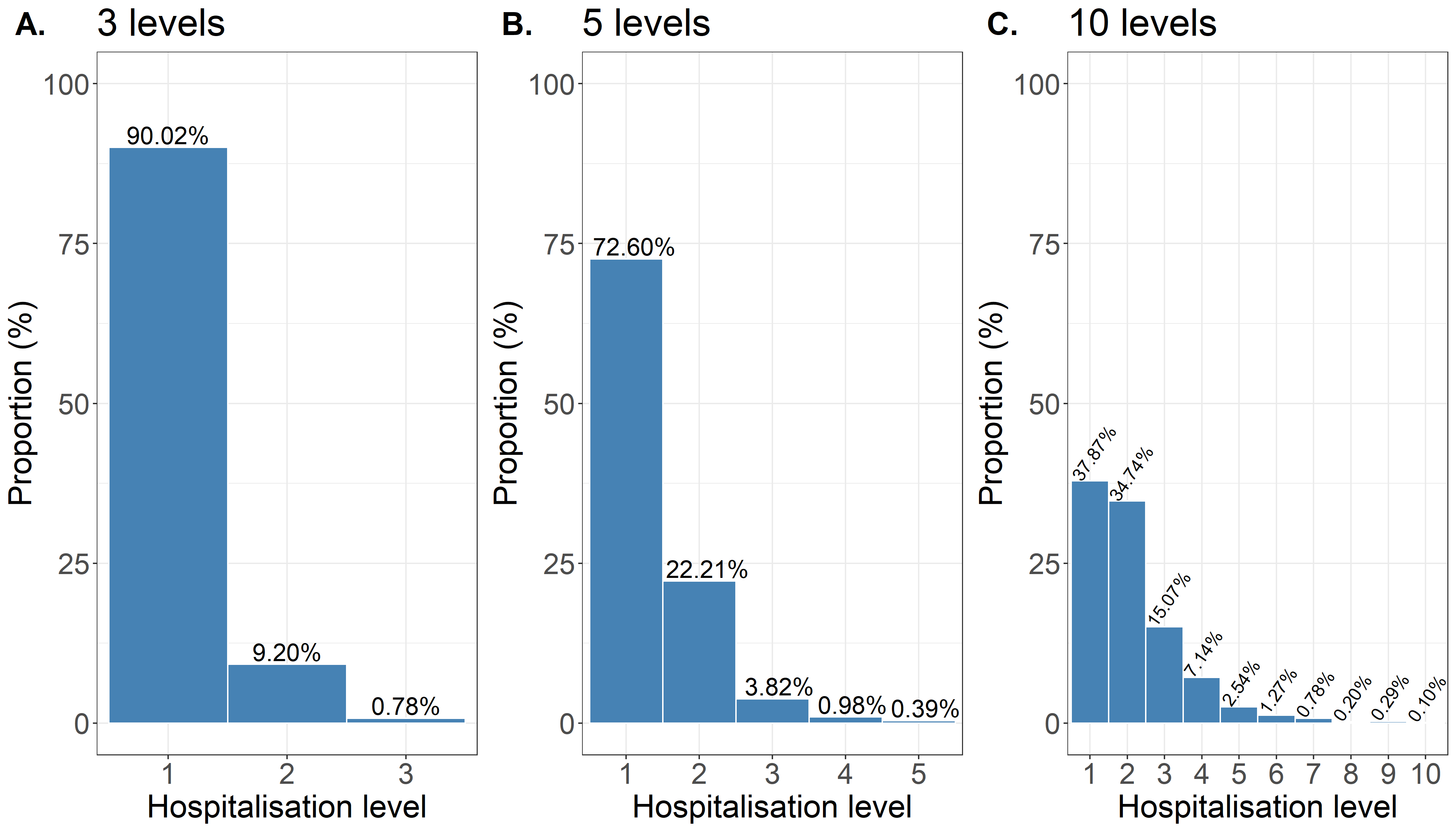

### S2A Figure.png

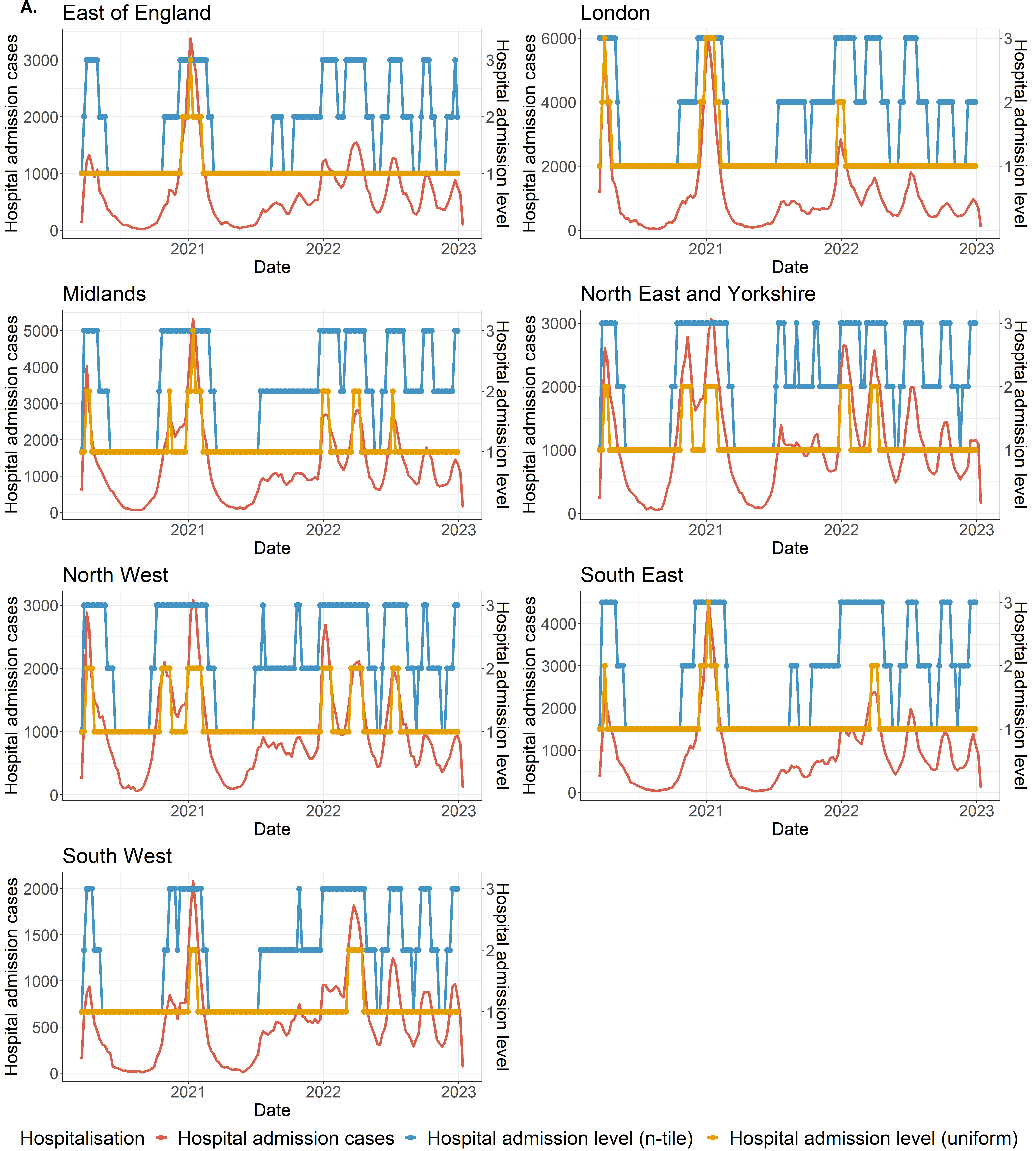

### S2B Figure.png

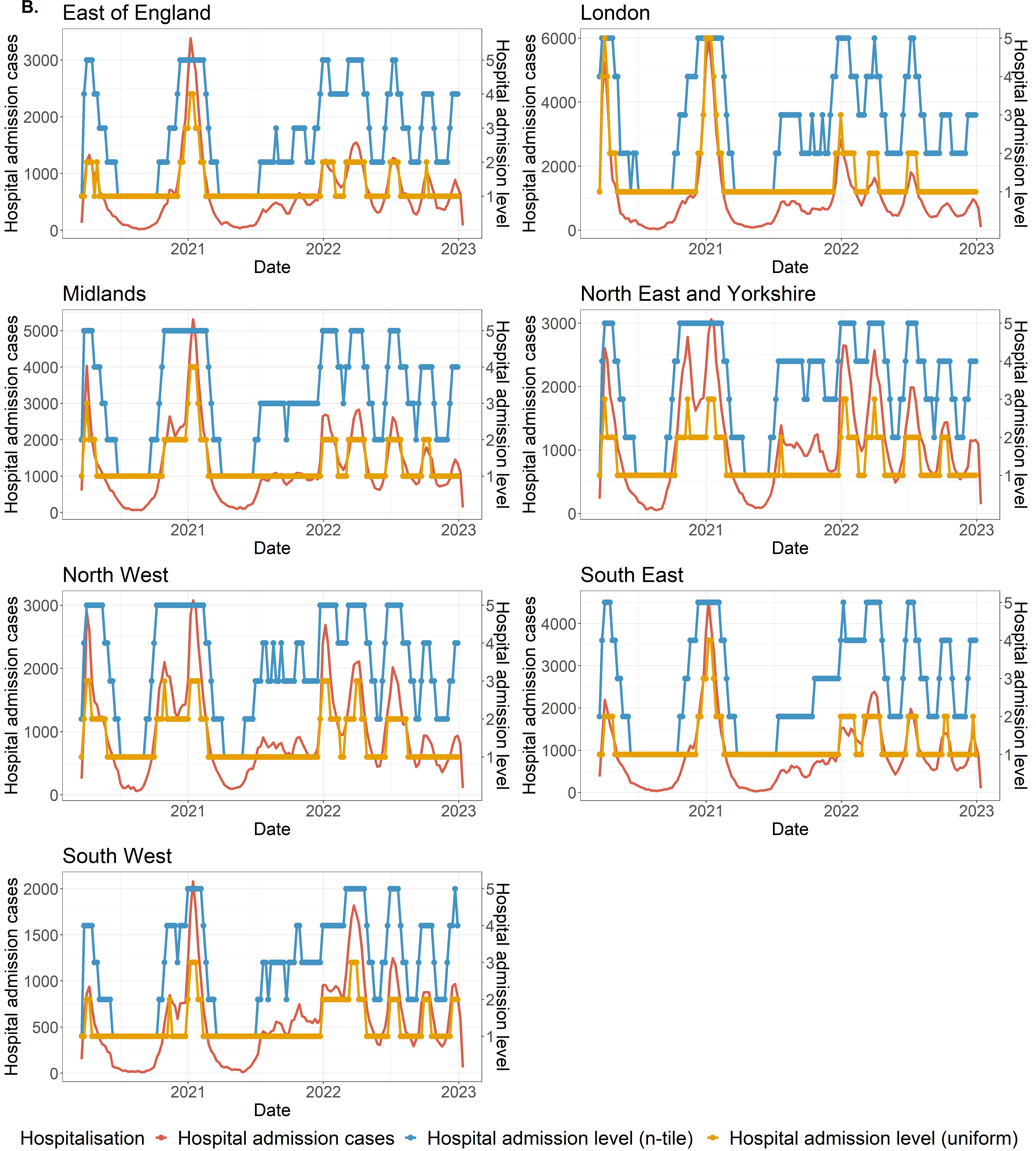

### S3 Figure.png

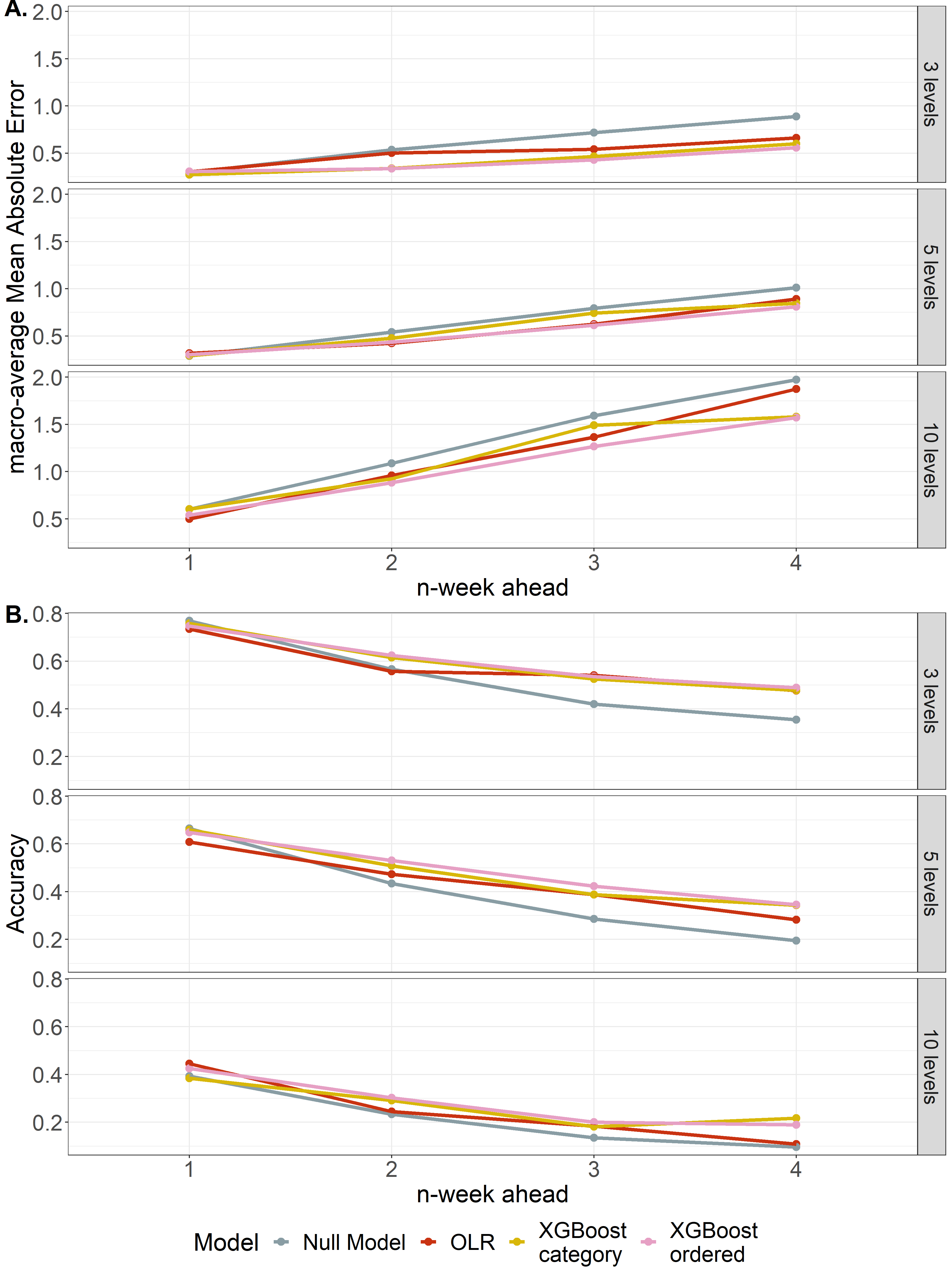

### S4 Figure.png

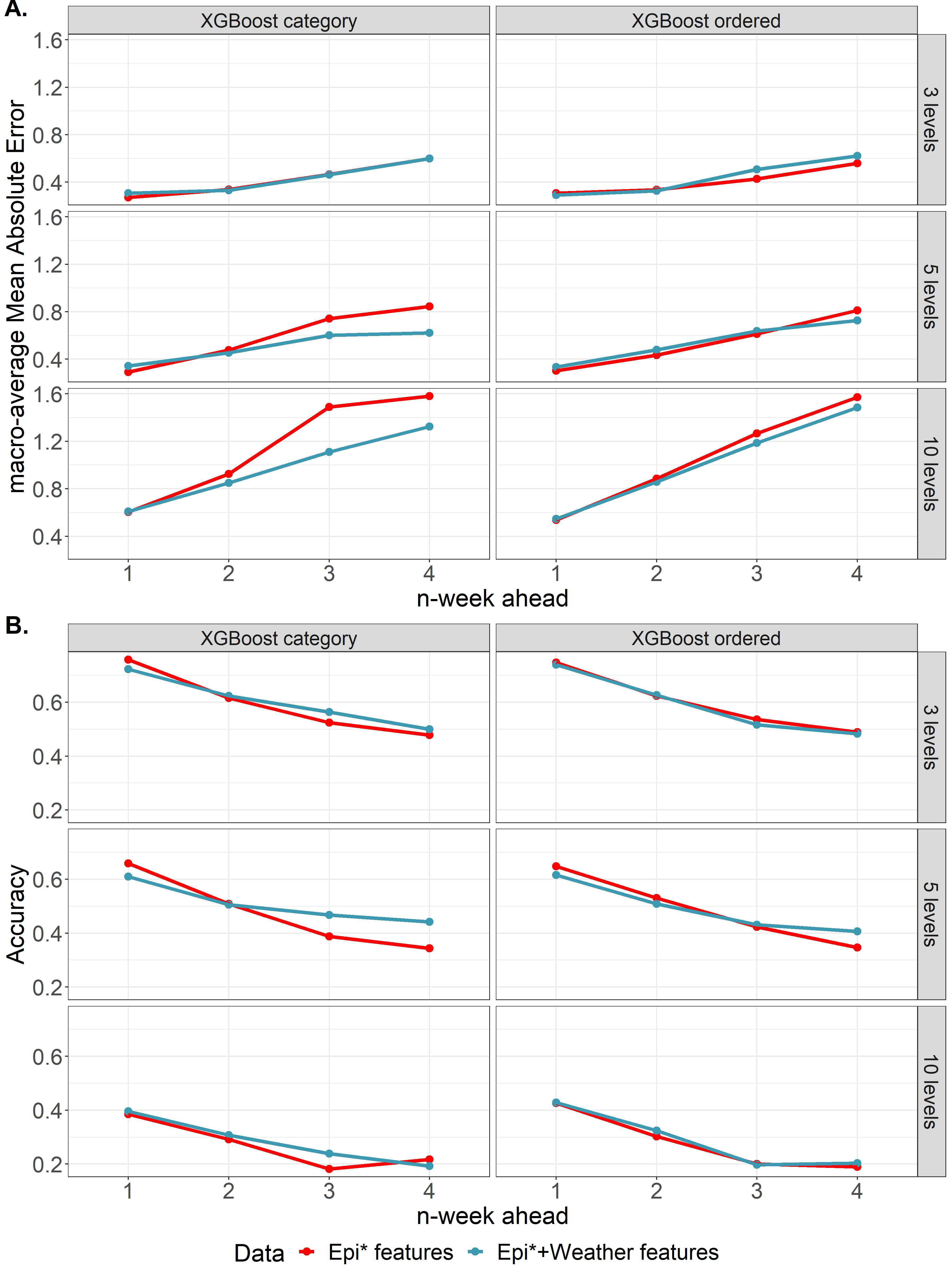

### S5 Figure.png

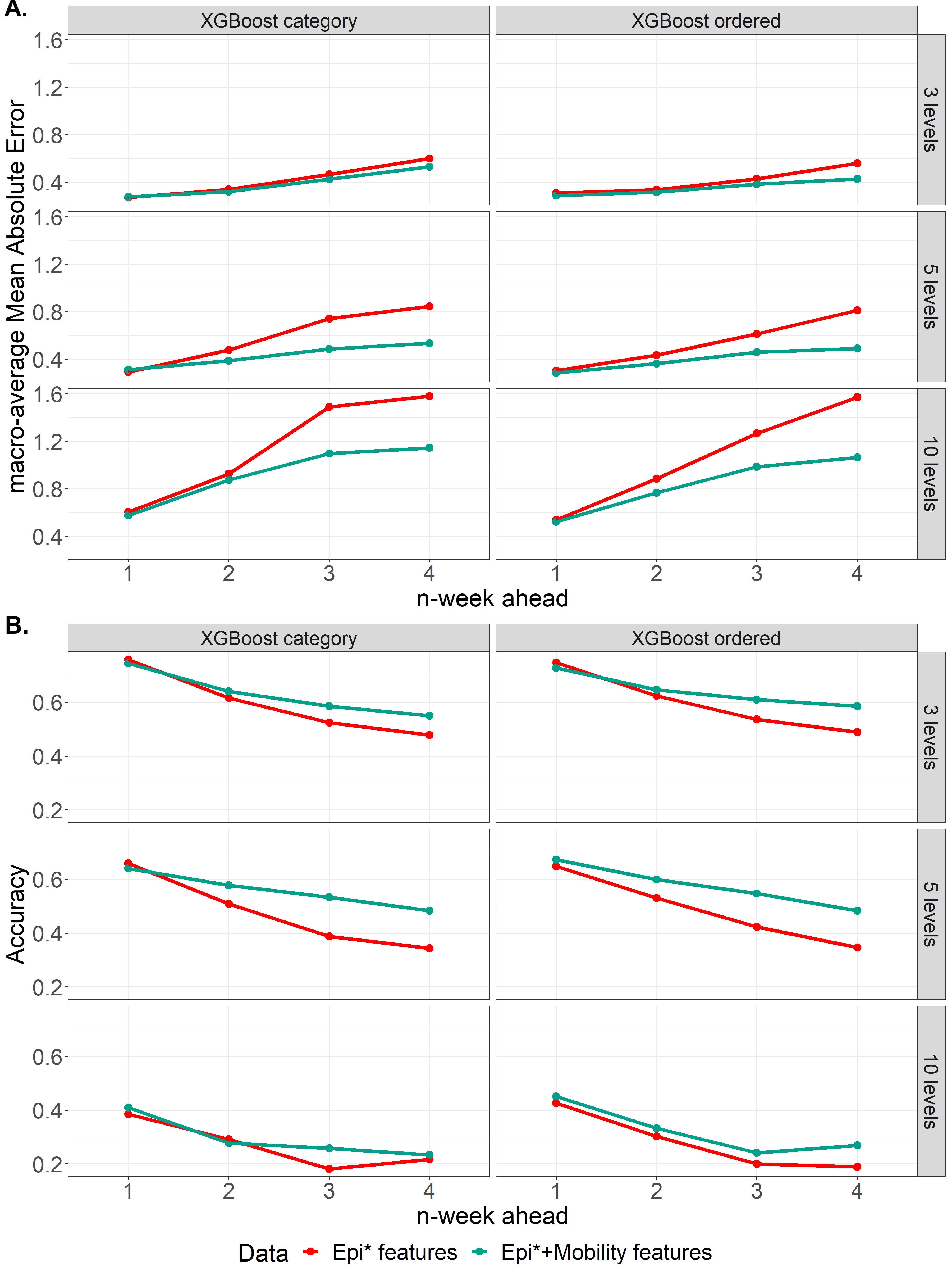

### S6 Figure.png

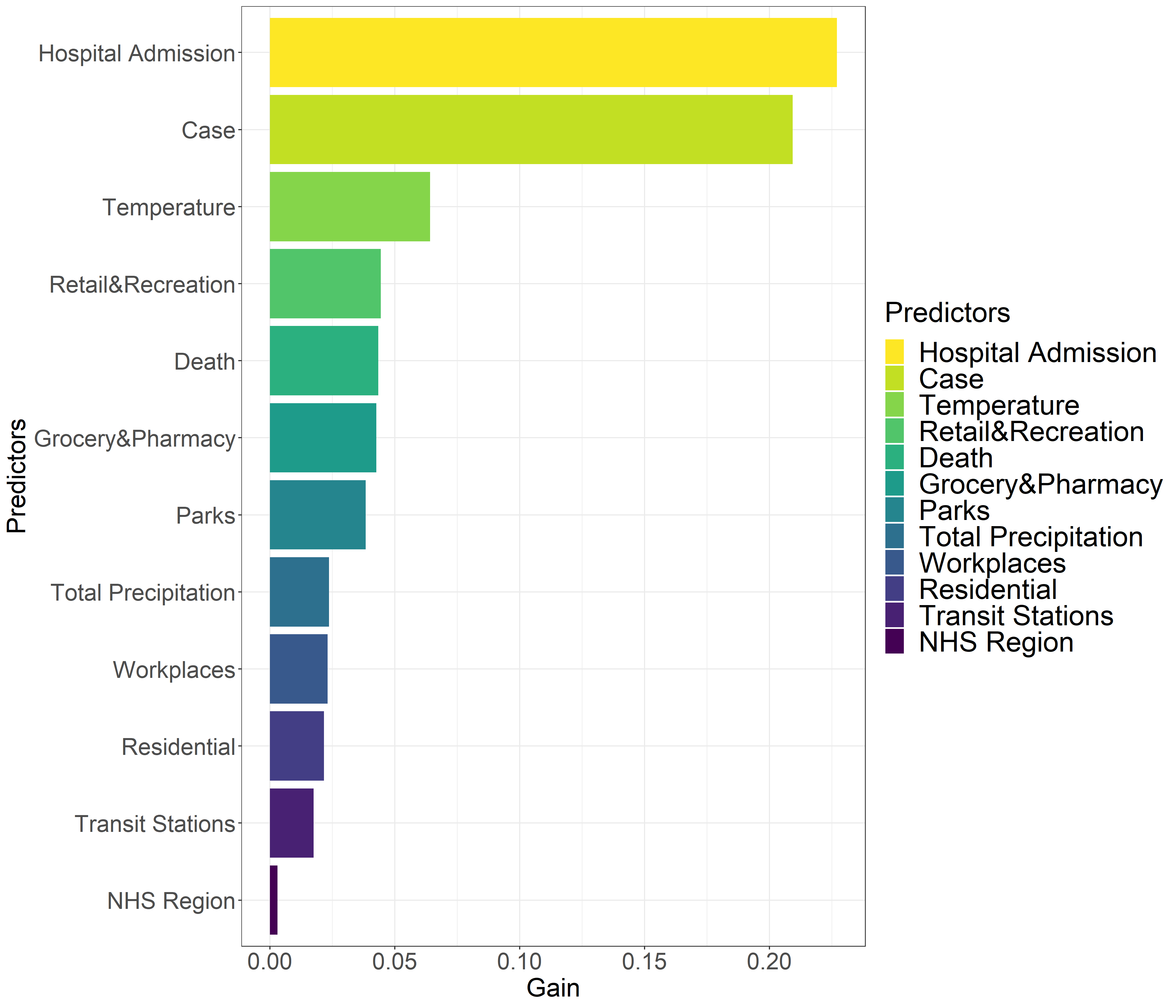
